# Dysconnectivity of a brain functional network was associated with blood inflammatory markers in depression

**DOI:** 10.1101/2021.04.02.21254853

**Authors:** Athina R. Aruldass, Manfred G. Kitzbichler, Sarah E. Morgan, Sol Lim, Mary-Ellen Lynall, Lorinda Turner, Petra Vertes, Wellcome Trust Consortium for Neuroimmunology of Mood Disorders and Alzheimer’s Disease (NIMA), Jonathan Cavanagh, Phil Cowen, Carmine M. Pariante, Neil A. Harrison, Edward T. Bullmore

## Abstract

**Objective:** There is increasing evidence for a subgroup of major depressive disorder (MDD) associated with heightened peripheral blood inflammatory markers. In this study, the authors sought to understand the mechanistic brain-immune axis in inflammation-linked depression by investigating associations between functional connectivity (FC) of brain networks and peripheral inflammation in depression.

**Methods:** Resting-state functional magnetic resonance imaging (fMRI) and peripheral blood immune marker data (C-reactive protein; CRP, interleukin-6; IL-6 and immune cells) were collected on N=46 healthy controls (HC; CRP ≤ 3mg/L) and N=83 cases of MDD, stratified further into low CRP (loCRP MDD; ≤ 3 mg/L; N=50) and high CRP (hiCRP MDD; > 3 mg/L; N=33). In a two-part analysis, network-based statistics (NBS) was firstly performed to ascertain FC differences via HC vs hiCRP MDD comparison. Association between this network of interconnected brain regions and peripheral CRP (N=83), IL-6 (N=72), neutrophils and CD4+ T-cells (N=36) were then examined in MDD cases only.

**Results:** Case-control NBS testing revealed a single network of abnormally attenuated FC in hiCRP MDD, chiefly comprising default mode network (DMN) and ventral attentional network (VA) coupled regions, anatomically connecting the insula/frontal-operculum and posterior cingulate cortex (PCC). Across all MDD cases, FC within the identified network scaled negatively with CRP, IL-6, and neutrophils.

**Conclusions:** The findings suggest that inflammation is associated with attenuation of functional connectivity within a brain network deemed critical for interoceptive signalling, e.g. accurate communication of peripheral bodily signals such as immune states to the brain, with implications for the etiology of inflammation-linked depression.

## INTRODUCTION

Subsuming a heterogeneous population of patients into diagnostic categories that are chiefly defined by syndromic and behavioral constructs, as opposed to biological discriminators, has been a persistent issue in classification of major depressive disorder (MDD; henceforth also referred to as depression) (1). In view of this, there has been growing interest in identifying a subgroup of MDD cases associated with blood biomarkers of peripheral inflammation (2; 3), so-called inflammation-linked depression.

Evidence for mechanistic links between the immune system and depression, first nucleated scientifically about 30 years ago, and has since become the foundation of the emerging field of immunopsychiatry (4; 5; 6; 7; 8). Meta-analyses of cross-sectional, case-control studies have demonstrated low-grade increases in peripheral C-reactive protein (CRP) and inflammatory cytokines, particularly interleukin 6 (IL-6), in MDD cases (9; 10; 11). In population samples, higher levels of CRP and IL-6 at baseline predicted increased risk of depression at follow-up, suggesting a causal role for inflammation in depression (12). Whilst there is a clear implication for inflammation in the pathoetiology of a subgroup of MDD cases, but what this mechanistically entails across the neuroimmune axis still remains elusive (13; 14). Functional magnetic resonance imaging (fMRI) is a potentially useful tool for investigation of inflammation- and depression-related changes in brain functional connectivity (15; 16).

In case-control fMRI studies of depression, functional connectivity (FC) abnormalities have been frequently reported with a focus on canonical resting state networks (RSNs) such as the default mode network (DMN), the ventral attentional network (VA), and the fronto-parietal control network (FP), each of which is associated with specific behavioral domains and/or classes of symptoms relevant to MDD. For example, abnormal connectivity within the DMN has been linked to rumination and negative self-referential thoughts (17; 18); whereas dysconnectivity of the VA network has been associated with impairments in emotion recognition and processing, apathy and anhedonia (19). Nonetheless, there are inconsistencies between individual studies. Both abnormal hypoconnectivity (reduced positive FC and increased negative FC), and abnormal hyperconnectivity (increased positive FC and reduced negative FC), have been reported for the DMN, FP and VA networks in depression (20).

There have been fewer fMRI studies of inflammation-related changes in resting state connectivity, with greater paucity in studies with clinical depression sample. However, a recent meta-analysis encompassing human experimental models of inflammation, clinical studies of hepatitis C patients receiving IFN*α* treatment, and observational studies of community samples with variable blood levels of CRP, reported that inflammation-related changes were co-localized to DMN, VA and limbic functional networks (21). Seminal studies have reported CRP-related differences in connectivity among depressed cases (rather than between cases and controls), with high CRP (inflamed depression) negatively correlated with seed-based analysis of cortico-striatal and cortico-amygdalar FC (22; 23; 24). Overall, there is emerging evidence that peripheral inflammation can perturb FC of brain networks that are known to be critical for emotional regulation (25; 26; 27).

Here, we investigated the relationships between depression, peripheral inflammation and whole-brain functional connectivity in two related analyses, complementary to our previous investigation on structural and FC differences in HC and all MDD cases, on the same imaging cohort (28). First, we used networkbased statistics (NBS) on functional connectome to test for network-level FC differences between high CRP MDD cases (hiCRP MDD; > 3 mg/L) compared to healthy controls (HC; ≤ 3 mg/L). Informed by prior reports of decreased FC associated with inflammation in MDD (22; 23; 24) and with MDD in case-control studies (20; 29), we tested one-tailed hypothesis that there is no set of interconnected edges (or connections) with attenuated FC in hiCRP MDD compared to HC; and this null hypothesis was refuted. A network of significantly attenuated connectivity linking mainly insular, cingulate and subcortical regions was identified. Second, we used this network as a mask to explore within-group relationship between FC measured in all MDD cases, and peripheral inflammation indexed by CRP, IL-6 and immune cell counts. We hypothesized that increased blood protein and cellular inflammatory markers would be negatively correlated with FC of this network.

## METHODS & MATERIALS

### Participants

Biomarkers for Depression (BioDep) was an observational case-control study conducted as part of the Wellcome Trust Neuroimmunology of Mood Disorders and Alzheimer’s disease (NIMA) Consortium. All procedures were approved by an independent national research ethics service (NRES) committee (NRES: East of England, Cambridge Central, UK; Reference: 15/EE/0092) and all participants provided written informed consent. All participants satisfied inclusion criteria, e.g. aged 25-50 years, and exclusion criteria, e.g. major medical inflammatory disorder or immuno-modulatory medication **(Supplemental Appendix 1 (SA1))**. All MDD cases screened positive for current depressive symptoms on the Structured Clinical Interview for DSM-5 Depressive Disorders (SCID), and had global Hamilton Rating Scale for Depression (HAM-D) score > 13 **(SA3-4)**. After initial telephone screening, potentially eligible participants attended an eligibility assessment **(Figure S1)**, including blood sampling for CRP assay, at one of 5 UK recruitment centres (Brighton, Cambridge, Glasgow, King’s College London (KCL), or Oxford). Eligible participants next attended one of 3 UK assessment centres (Cambridge, KCL or Oxford) for venous blood sampling, clinical assessment, and MRI scanning, all scheduled on the same day **(SA2)**. MDD cases were then stratified by blood CRP level: loCRP MDD had CRP ≤ 3mg/L, hiCRP MDD had CRP > 3mg/L. All HC had CRP ≤ 3mg/L.

### Blood immune biomarkers

All participants provided up to 90mL of fasting venous blood. CRP was measured using high sensitivity immunoturbidometry at a central laboratory (Q^2^ Solutions, UK). Cytokine and chemokine levels were measured in plasma and serum using relevant V-PLEX 10-spot immunoassay kits from Meso Scale Discovery (MSD) **(see Section S2)**. After quality control (QC), analyzable cytokine data on IL-6 and other cytokines were obtained for N=72 MDD cases (**?**). However, only IL-6 was analyzed further in the present study as majority of evidence linking depression to peripheral inflammation show more reproducible association between CRP and IL-6 with depression (30; 31; 32). All concentrations were log-transformed (base 10).

### Cellular biomarkers

Absolute cell counts were available for 12 leukocyte classes for N=36 MDD cases **(Table S1B-C)** that were a subset of a previous report on a larger sample (11). We then used binary classification outcome from this prior study defined using 2-way forced clustering analysis using multivariate mixture modelling on cellcounts – to assign each MDD sample in the current study into inflamed or uninflamed -MDD subgroups **(Figure S3)**. We recognize that in the context of systemic inflammation in depression, some immune cells are perhaps more relevant for scrutiny than others. Hence, for the purpose of this study, we examined neutrophils and CD4+ (helper) T-cells, since these are the principal effectors of innate and adaptive immune responses respectively (33; 34) and were significantly increased in MDD subgroups as reported in a prior study (11).

### Functional magnetic resonance imaging

Resting-state fMRI data were acquired using multi-echo (me) echoplanar imaging (EPI) sequence with the following parameters: relaxation time (TR)=2.57s; echo times (TE_1_,_2_, _3_)=15ms, 34ms and 54ms; acquisition time = 10mins 42.5s = 250 time points in each fMRI time series. meEPI data were collected as 32 slices at −30 degrees to the AC–PC line with field of view=240mm and matrix size = 64 x 64, for voxel resolution of 3.75 x 3.75 x 3.99mm. The first six volumes were discarded and remaining data preprocessed using multi-echo independent component analysis (ME-ICA) (35; 36) in AFNI. Images were then regionally parcellated using a 180 bilateral cortical surface–based atlas (37) and 8 bilateral non–cortical regions per FreeSurfer (38; 39), resulting in a 376 x 244 regional timeseries. Timeseries were then bandpass filtered at wavelet scales 2 and 3 corresponding to 0.02–0.1Hz. The FC between each regional pair was estimated by Pearson’s correlation coefficient r between pairwise wavelet coefficients and then averaged, resulting in a 376 x 376 symmetric FC matrix. FC matrix were then Fisher r-to-z transformed. Subjects with high degree of head motion estimated by framewise displacement, FD_*max*_ > 1.3mm and/or FD_*rms*_ > 0.3mm were excluded (N=4). Additional nuisance variables i.e. FD_*rms*_, scan site and age were regressed edge–wise from the FC matrices **(Figure S4)**.

### Part 1 analyses: Network–based statistics (NBS) and group difference in network connectivity

NBS was implemented using the NBS MATLAB Toolbox (40) **(see Section 4 Supplemental Data)**. We performed NBS case-control comparison on HC vs hiCRP MDD. We used one-tailed t-tests (HC > hiCRP MDD) and performed 5000 random permutations. The test statistic threshold was initially set to t_*primary*_=3.0, corresponding to the nominal uncorrected *P*=0.005 and was reported to yield consistent findings across parcellation schemes (41; 42). It was then increased by 0.1 step-wise, to retain edges with strongest experimental effect i.e. greatest t-statistic, at the same time generating a statistically significant component. NBS results at t_*primary*_=3.8 were reported here and presented as three-dimensional network visualizations using BrainNet Viewer (43).This output was examined categorically in all three groups and subsequently used as a “mask” for Part 2 analyses with immune markers.

### Part 2 analyses: Association between functional connectivity and blood immune biomarkers

In the second part of our investigation, we examined average network connectivity estimated from the HC vs hiCRP MDD NBS mask from Part 1 against CRP, IL-6, neutrophils and CD4+ T-cells, in all MDD cases only. This planned two-stage analysis avoids circularity by using a different pool of sample for the different stages of analysis.

### Statistical analysis

Case-control and within-group comparisons of FC distribution were estimated using two-sample KolmogorovSmirnov tests. Correlation between edge-wise FC within selected NBS network and immune markers was estimated using Pearson’s correlation. Sensitivity analyses were performed using hierarchical linear regression to control for the additive effects of covariates including clinical and diagnostic variables, sociodemographic and lifestyle variables, and behavioural measures **(Table S3B-C)**. Functional connectivity estimated by inter-regional time series correlated were normalised by Fisher’s r-to-*Z* transformation prior to analyses. Effect sizes were also reported using Cohen’s d.

## RESULTS

### Sample characteristics

After QC procedures, analysable fMRI and CRP data were available for N=129 participants, comprising N=46 HC, N=50 loCRP MDD and N=33 hiCRP MDD. Sociodemographic and clinical variables are summarised in Table 1. hiCRP MDD included proportionally more females and had higher BMI and more severe depression scores than loCRP MDD. BMI was also significantly correlated with CRP (r=0.57, *P*_*FDR*_= 0.001) and IL-6 (r=0.45, *P*_*FDR*_ < 0.05). Therefore, sex and BMI were included as covariate in NBS testing and subsequent statistical model **(Table S1A-C)**. Depressed cases were eligible for participation if currently prescribed antidepressants and pre-specified concomitant medication for minor diagnostic comorbidities (see **SA4** for details). Potential confounding effects of antidepressant and other medication and clinical comorbidities, as well as sex and BMI, were controlled for as part of sensitivity analyses.

**Table 1:** Sociodemographic characteristics, clinical and serological variables in the analyzable cohort N=129). Group difference were estimated using Mann-Whitney U test or chi-squared test ^*a*^body mass index.1 HC missing data omitted in case-control statistical comparison computation; ^*b*^Hamilton Rating Scale for Depression; ^*c*^Beck’s Depression Inventory (version II); ^*d*^Snaith-Hamilton Pleasure Scale; ^*e*^Chalder Fatigue Scale; ^*f*^ State-Trait Anxiety Inventory; ^*g*^Childhood Trauma Questionnaire; ^*h*^Perceived Stress Scale; ^*i*^Life Events Questionnaire; ^*j*^high-sensitivity C-reactive protein. HC; healthy controls (CRP ≤ 3mg/L); all MDD; all MDD cases (CRP 3–10 mg/L); loCRP MDD; low CRP MDD cases (CRP ≤ 3mg/L); hiCRP MDD; high CRP MDD cases (CRP > 3mg/L); IQR; interquartile range (Q3 – Q1); ^†^statistical comparison performed using unpaired t-test; ^∗^p < .05; ^∗∗^p < .01; ^∗∗∗^p < .001.

|  | HC<br>(N=46) |  | all MDD<br>(N=83) |  | Case-Control<br>difference | loCRP MDD<br>(N=50) |  | hiCRP MDD<br>(N=33) |  | loCRP–hiCRP<br>difference |
| --- | --- | --- | --- | --- | --- | --- | --- | --- | --- | --- |
|  | Median | IQR | Median | IQR | p-value | Median | IQR | Median | IQR | p-value |
| **Sociodemographic / Clinical** |  |  |  |  |  |  |  |  |  |  |
| Age (years) | 34.5 | 11.9 | 36.3 | 12.6 | 0.21 | 37.6 | 11.4 | 36.3 | 12.6 | 0.54 |
| BMI (kg/m<sup>2</sup>)<sup>a</sup> | 23.5 | 5.3 | 26.6 | 5.9 | **0.001\*\*** | 25.4 | 4.6 | 27.8 | 7.4 | **0.001\*\*** |
| Sex, Male (n, %) | 19 | 41.3 | 26 | 30 | 0.34 | 21 | 43.8 | 5 | 16.7 | **0.02\*** |
| Tobacco, current smokers (n, %) | 2 | 4.3 | 15 | 19.2 | **0.05\*** | 11 | 22.9 | 4 | 13.3 | 0.39 |
| Alcohol, current users (n, %) | 23 | 50 | 47 | 60.3 | 0.59 | 30 | 60 | 17 | 51.5 | 0.59 |
| Cannabis, current users (n, %) | 4 | 8.7 | 5 | 6.4 | 0.95 | 4 | 8 | 1 | 3 | 0.65 |
| Handedness, Right (n, %) | 42 | 91.3 | 69 | 88.5 | 0.29 | 41 | 82 | 28 | 84.8 | 0.94 |
| Ethnicity, White (n, %) | 31 | 67.4 | 67 | 85.9 | 0.55 | 43 | 86 | 24 | 72.7 | 0.24 |
| **Behavioral Instruments** |  |  |  |  |  |  |  |  |  |  |
| HAM–D (17–items)<sup>b</sup> | 0 | 1 | 17 | 7 | < **0.001\*\*\*** | 17 | 7 | 18 | 5 | 0.44 |
| BDI–II<sup>c</sup> | 1 | 3 | 25 | 11.5 | < **0.001\*\*\*** | 25 | 10.8 | 26 | 10 | 0.56 |
| SHAPS<sup>d</sup> | 0 | 0 | 4 | 5.5 | < **0.001\*\*\*** | 5 | 4 | 3 | 6 | 0.22 |
| CFS<sup>e</sup> | 11 | 1 | 20 | 7 | < **0.001\*\*\*** | 20.5 | 7 | 20 | 6 | 0.94 |
| STAI – S (items 1 – 20)<sup>f</sup> | 25 | 8.8 | 51 | 15 | < **0.001\*\*\*** | 52.5 | 15.5 | 49 | 12 | 0.63 |
| STAI – T (items 21 – 40)<sup>f</sup> | 28.5 | 9 | 61 | 11.5 | < **0.001\*\*\*** | 61 | 11.8 | 61 | 12 | 0.95 |
| CTQ<sup>g</sup> | 37 | 5.8 | 51 | 22 | < **0.001\*\*\*** | 53 | 20.8 | 47 | 16 | 0.06 |
| PSS<sup>h</sup> | 11.5 | 8 | 26 | 6 | < **0.001\*\*\*** | 26.5 | 5.6 | 26 | 6 | 0.73 |
| LEQ (score)<sup>i</sup> | 0 | 1 | 1 | 1 | < **0.001\*\*\*** | 1 | 2 | 1 | 1 | 0.61 |
| LEQ (rating)<sup>i</sup> | 1.5 | 1 | 3 | 4 | < **0.001\*\*\*** | 3 | 4 | 3 | 3.5 | 0.91 |
| **Inflammatory Biomarkers** |  |  |  |  |  |  |  |  |  |  |
| CRP (mg/ L)<sup>j</sup> | 0.6 | 1.1 | 1.8 | 3.5 | < **0.001\*\*\*** | 0.8 | 1.0 | 4.5 | 3.7 | < **0.001\*\*\*** |
| CRP (log<sub>10</sub> mg/L)<sup>j†</sup> | -0.2 | (0.4) | 0.2 | (0.5) | < **0.001\*\*\*** | -0.1 | (0.4) | 0.7 | (0.2) | < **0.001\*\*\*** |

### Case–control network-level differences in FC

one-tailed NBS comparison between hiCRP MDD and HC resulted in a single network comprising 38 edges and 33 nodes (or regions) (one-tailed *P*=0.043, Cohen’s d=0.45) **(Figure 1A)**. The nodes of this network were affiliated to VA and DMN cortical areas and subcortical nuclei, based on Yeo-7 functional network assignment (each network referred to as “module”) (44). Edges between DMN and VA nodes (10/38) were the most abundant intermodule connections, followed by DMN-Somatomotor (8/38) edges. Anatomically, edges were mainly coupled between insular/frontal-opercular cortex and PCC, and abutting parietal cortical areas with limbic area e.g. ACC, thalamus, and putamen **(Table S2, Figure S5B)**. Controlling for sex yielded single significant network with strictly DMN-VA edges, with greater effect size (highest network yielding primary threshold t_*primary*_=9.0, *P*= < 0.001, Cohen’s d=0.52; Figure S5B). Controlling for sex and BMI resulted in no significant network.

**Figure 1:**
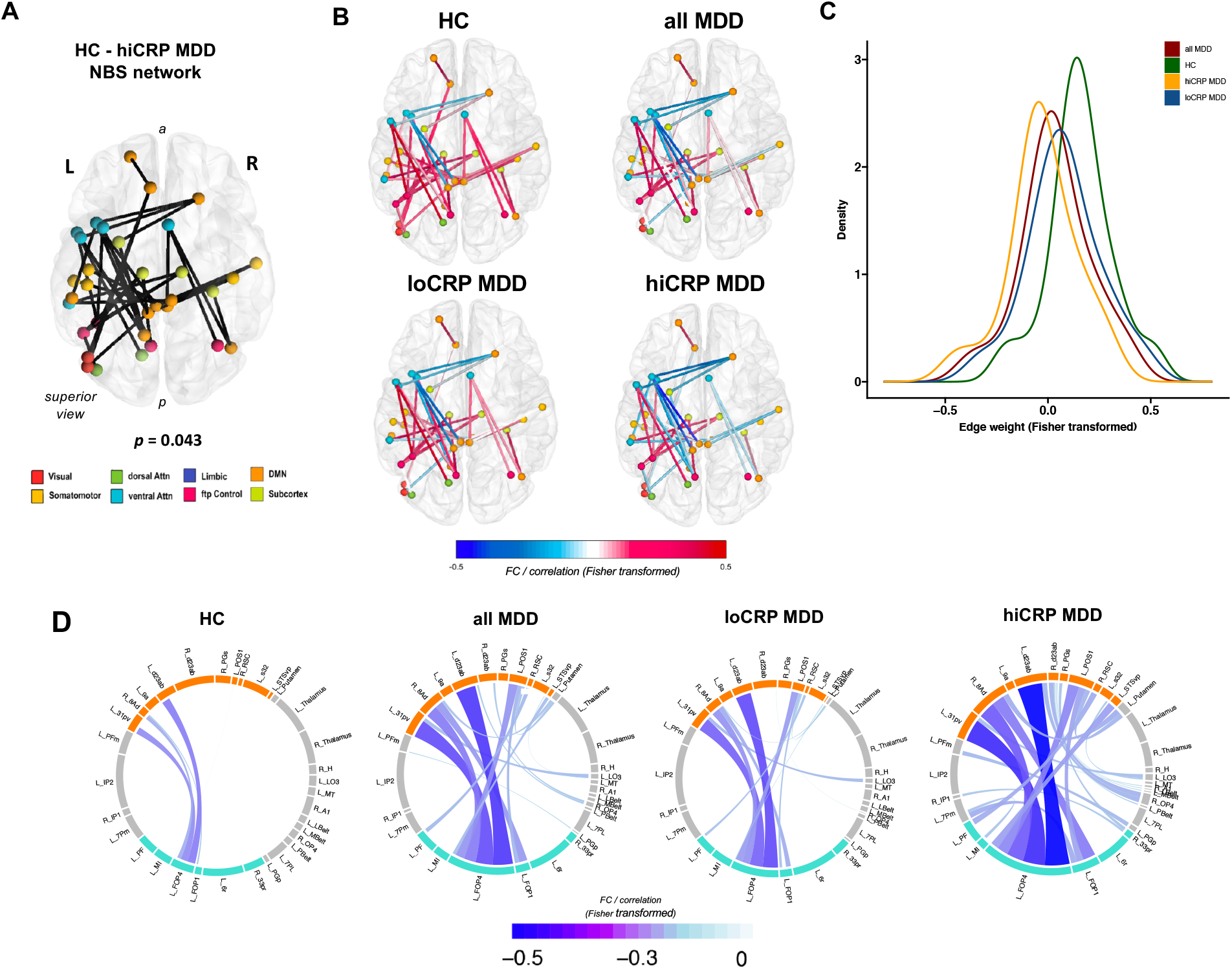
Network-based case-control differences in functional connectivity. **(A)** Network of case-control (HC vs hiCRP MDD) FC differences – modelling composite effect of inflammation *and* depression – identified by Network Based Statistics (NBS). A network comprising 38 edges or connections, across 33 nodes or regions, was generated by testing a one-tailed hypothesis, i.e. HC > hiCRP MDD, and reported here at the highest primary threshold t_*primary*_=3.8, at which a significant (*P*¡0.05) network was identifiable. **(B)** Group-averaged functional connectivity across this case-control network, plotted for each group. Across HC, loCRP MDD and hiCRP MDD, reduced functional connectivity (FC) was observed (more blue edges). These edges were mainly coupling default mode network (DMN)-ventral attentional (VA) functional modules, and anatomically, the insular/frontaloperculum and posterior cingulate cortical regions. **(C)** Distribution of edge-weights within the HC vs hiCRP MDD NBS network per group. **(D)** Distribution of negative edges within the HC vs hiCRP MDD NBS network across groups, visualized in topological space via radial network diagram. The outer track is divided into sectors, each denoting a node within the NBS network. Width of the sector denotes approximate weighted nodal degree i.e. the greater the number of (negative) connections, the longer the sector. Thickness and colour of links denote the strength of functional connectivity (FC), where edges with more negative FC have thicker and bluer links. Links within each sector are ordered clockwise with increasing FC. Across HC, loCRP MDD and hiCRP MDD groups, increases in both number and weight of negative edges were observed, especially between the DMN and VA functional modules.

FC over all edges network was less negative in HC (−0.22 < *Z* < 0.51), than in loCRP MDD (−0.35 < *Z* < 0.46) and hiCRP MDD MDD (−0.46 < *Z* < 0.31) **(Figure 1B-C)**. Distribution of edge-weight was significantly different between hiCRP MDD vs HC (KS test, *P*=1.6 × 10^−12^, D=0.61, Cohen’s d=1.13), loCRP MDD vs HC (KS test; *P*=0.00027, D=0.34, Cohen’s d=0.47) and hiCRP MDD vs loCRP MDD (KS test; *P*=0.00027, D=0.34, Cohen’s d=0.61). Topological representation of the network confirmed that edges with attenuated FC were mainly between DMN-VA functional networks, with common set of edges showing lowest correlation across groups i.e L FOP4–L d23ab, L FOP4–L 31pv, L FOP4–R 8Ad **(Figure 1D)**. ^1^

To theoretically define this network, we last queried the BrainMap database (45) to identify fMRI studies that showed co-activation for both insula and cingulate cortices. The 8 resultant studies **(Table S2C)** involved subjective perception of various stimuli, self-referential and salience processing, all of which are interoceptive processes (46).

### Association between FC and inflammatory proteins in MDD

We next examined the relationships between inflammatory proteins and edge-wise FC within the mask defined by HC vs hiCRP MDD NBS network, in all MDD cases. Both IL-6 (−0.49 < *Z* < 0) and CRP (−0.48 < *Z* < 0) were negatively correlated with edges within the network **(Figure 2A, first column)**. When testing each edge separately for association with each inflammatory protein, while controlling for multiple comparisons with FDR<5%, 21 edges were significantly negatively correlated with CRP and 6 edges with IL-6 **(Figure 2A, second column; Table S3A)**. These 6 edges were a subset of the 21 edges significantly negatively correlated with CRP and were mainly the previously highlighted DMN-VA intermodular edges which were anatomically localised between insular/frontal-opercular cortex and PCC **(Figure 2B)**.

**Figure 2:**
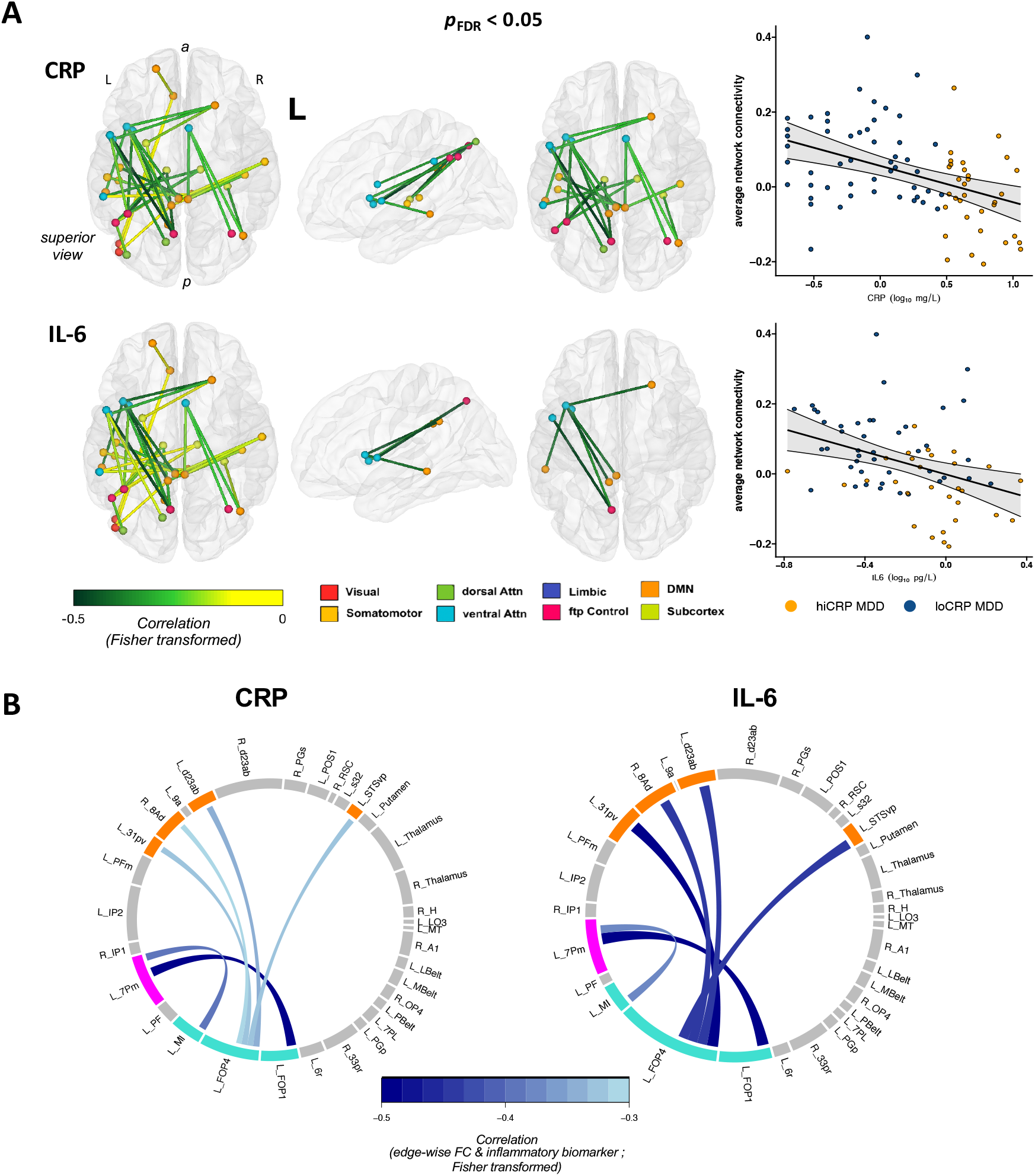
Functional connectivity association with inflammatory proteins in all MDD cases. **(A**) First column: edge-wise functional connectivity (FC) within HC vs hiCRP MDD network correlated with CRP (N=83 MDD) and IL-6 (N=72 MDD). Second column: : a subset of edges had functional connectivity significantly correlated with inflammatory protein concentrations (FDR < 5%). Third column: scatterplots of the continuous relationships between average network connectivity and blood concentrations of CRP and IL-6 with the best-fitting regression lines shown with 95% confidence intervals band. **(B)** Strength of correlation between edge-wise FC and blood concentrations of CRP and IL-6 are denoted by intensity of blue colouration and link thickness. Only edges with attenuated connectivity significantly related to both CRP and IL-6, surviving FDR correction are shown here. IL-6 appears to show more robust negative correlation with FC compared to CRP across all edges.

We then averaged FC over all edges within NBS mask to investigate how individual differences in FC within this network were related to inflammation. Average network connectivity (ANC) negatively correlated with CRP (r= −0.41, *P*=0.00008) and IL-6 (r= −0.36, *P*=0.0013) **(Figure 2A, third column; Table S3B)**. Associations remained significant after sex and BMI adjustment, where effects were generally more robust after covariate adjustment **(Table S3B)**. We also performed an additional sensitivity analysis to test the specificity of the observed relationship between ANC and inflammatory proteins to the NBS threshold (t_*primary*_=3.8). The correlation was robust across three other primary thresholds (t_*primary*_=3.1, 3.3, 3.5) **(Figure S6A-B)**.

### Association between FC and cellular markers in MDD

We finally sought to investigate difference in FC within NBS mask in the cell-stratified MDD subgroups. Analyzable cell count data were available on N=36 MDD cases, where neutrophils showed significant correlation with CRP (r=0.57, *P*_*FDR*_ < 0.001) and IL-6 (r=0.47,*P*_*FDR*_ < 0.05) **(Figure 3A, Table S1B)**. MDD subgroups significantly differed in CRP, basophils, eosinophils, neutrophils, classical monocytes, intermediate monocytes and NK-Tcells **(Table S1C)**. The “inflamed-MDD” subgroup had significantly lower FC than the “uninflamed-MDD” subgroup for the NBS network (KS test, *P*=2.2 x 10-16, D=0.11, Cohen’s d=0.12) **(Figure 3B)**. Consistent with previous observations, negative edges were again concentrated between DMN-VA modules, with identical edges showing greatest negative correlation **(Figure 3C)**. Neutrophils were negatively correlated (−0.41 ¡ *Z* ¡ 0) with edges within NBS, showing similar pattern of association to CRP and IL-6, although no individual edges demonstrated significant association with neutrophils after FDR correction. Neutrophils also scaled negatively (r= −0.34, *P*=0.025, *P*_*FDR*_ = 0.05) against ANC. We observed a similar trend for CD4+ T-cells, although concentrations were only weakly associated with edge-wise FC **(Figure S7; Table S3B)**.

**Figure 3:**
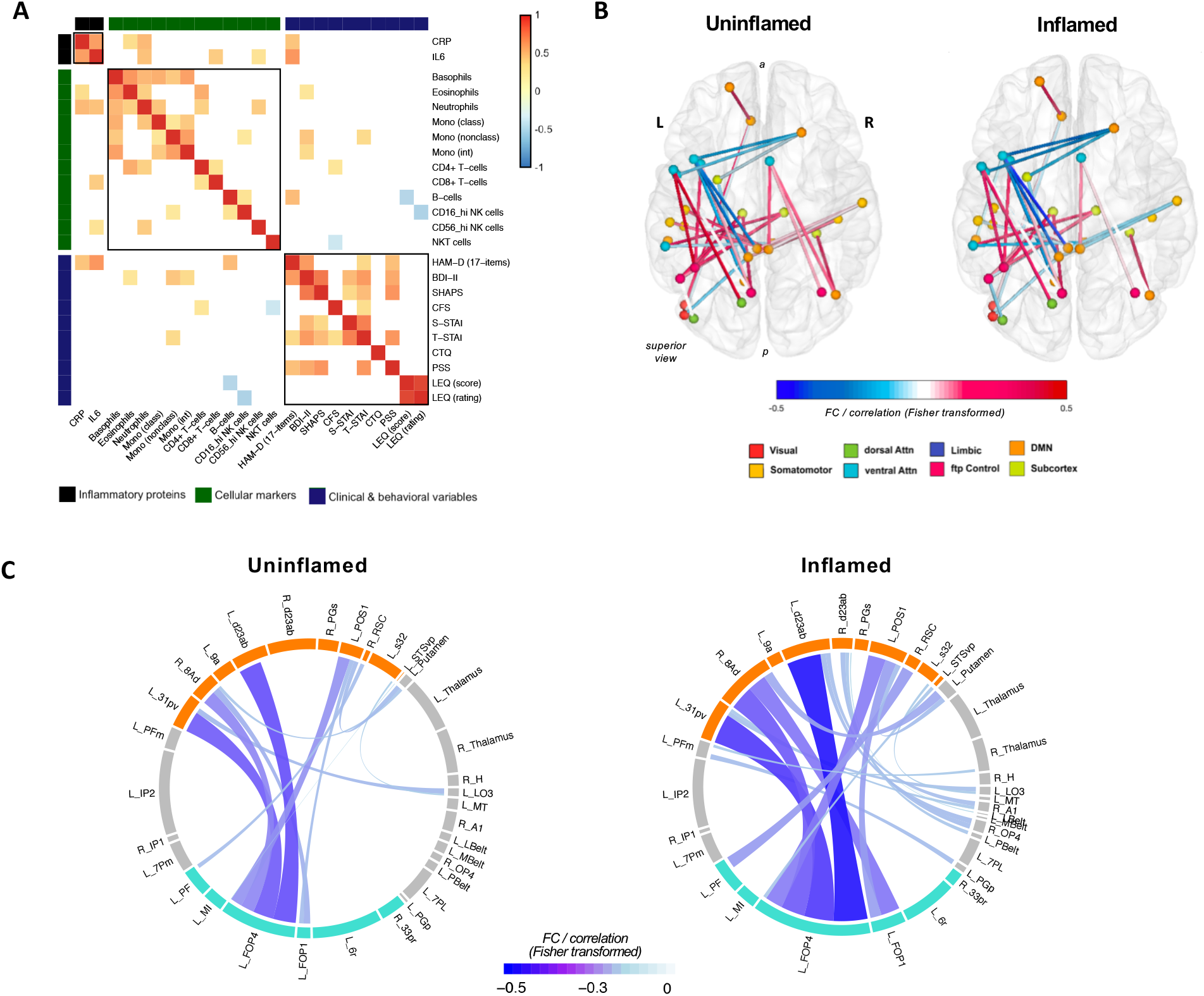
Functional connectivity differences between inflamed and uninflamed -MDD subgroups defined by immune cell counts. **(A)** Heatmap highlighting significant correlation between inflammatory proteins, cellular markers and clinical variables thresholded at *P* < 0.05 (N=36, all MDD). **(B)** Group-averaged representation of HC vs hiCRP MDD NBS network. **(C)** Distribution of negative edges within the NBS network comparing immune cell-stratified inflamed and uninflamed-MDD subgroups reported in Lynall et al. (2020). The inflamed-MDD subgroup shows a greater number of more negative edges, denoted by thicker and bluer links.

## DISCUSSION

As predicted by the first hypothesis, we found a network of interconnected edges with significantly reduced FC in MDD cases with heightened peripheral inflammation compared to HC. This NBS-derived network comprised edges localized primarily to connections between DMN and VA functional networks, linking the left insular/frontal-opercular and left PCC cortical regions. Group comparison of network connectivity revealed a hierarchical increase in FC attenuation, with HC showing least impairment, followed by loCRP MDD, and then hiCRP MDD. These NBS findings corroborate and extend outcomes from our earlier study which analysed between-group FC differences on the same dataset (28).

Next, we demonstrated negative scaling between CRP, IL-6, and average network FC, within the MDD cases only, supporting our second hypothesis. Our analyses using cell-stratified assignments, corroborated this observation in that more negative connections were noted in inflamed compared to uninflamed MDD subgroups, implicating identical connections and edges to those stratified by CRP.

### Interoceptive network dysfunction in inflammation-linked depression

More broadly,these results were consistent with evidence suggesting interoceptive dysfunction in inflammation-linked depression (47; 27). Interoception is the perception of bodily physiological states such as cardiovascular, gastrointestinal, pain, immune, and autonomic systems, that have been deemed as sources of emotional experience (48). Interoceptive processing, signalling and awareness i.e. the ability to “feel” what is happening within the body, is critical for emotional regulation, bodily homeostatic functioning and survival (48). In idiopathic MDD, interoceptive dysfunction has been associated with reduced emotional experience i.e. “feeling nothing”, alexithymia and anhedonia (49). In inflammation-linked depression, the interoceptive model provides some mechanistic insight by positing a role for bidirectional brain immune-signalling via neural pathway (predominantly the vagus nerve), that underpins communication of peripheral immune states to the brain and vice versa (6; 27). Collectively described as the “interoceptive nervous system” (INS), the INS is thought to relay afferent vagal information from the medulla, to a brain system anchored in the anterior insular, opercular, cingulate and somatomotor cortices (46; 48; 49; 50; 51). Regions within the INS are largely native to two canonical functional networks, namely the DMN and VA (also known as the salience network or cingulo-opercular network) (21; 52; 53). We therefore interpreted our NBS-derived network of attenuated FC in inflammation-linked depression, as indicative of disrupted function of the INS.

Concept of dysconnectivity in interoceptive systems has been previously evidenced by human experimental models of inflammation-linked depression i.e. “sickness behaviour” following immune challenge. In particular, Dipasquale et al. (2016) applied NBS to investigate FC differences preand post-IFN therapy in hepatitis-C positive participants (54). This study identified a similar network comprising bilateral insula, frontal cortex and subcortical regions, showing reduced FC 4-hours after IFNα induction in participants. Interoceptive signalling has also been described to show domain specificity and hierarchical processing. Thus, higher-level neural processing of interoceptive signals could vary between signal types e.g. affective (emotion), visceral physiology (immune, cardiac) or nociceptive (pain) inputs (47; 55). As such, subsystems of the INS may exist. For example, interoception is also linked to motivational circuit and could modulate reward sensitivity and hedonic sensing (26; 56). Attenuation within the corticostriatal and cortico-amygdalar pathways of the reward system has been reported with increased CRP in MDD (21; 22). A different study on the same dataset, also demonstrated using PBNA (parcellation-based network-analysis) – connectome-based technique similar to NBS (but using Bayesian multilevel modelling) – by seeding a cluster of vmPFC voxels, that increased CRP was again associated with decreased FC between the seed-cluster and a network of cortical areas comprising classic INS regions e.g. insula and cingulate cortex (24). These prior reports are broadly convergent with our suggestion that “inflammation-sensitive” brain regions such as the insula and striatum are likely embedded within a subsystem of the INS critical for parsing immune and affective signals.

### Interoceptive immune-sensing dysfunction in inflammation-linked depression

Findings from our secondary analyses, although less precedented, were also consistent with prior evidence. The negative scaling of FC against blood inflammatory markers corroborated reports from clinical MDD and population studies using a priori defined regions. In the former, heightened peripheral CRP, IL6, IL-1β and IL-1Ra negatively covaried with vmPFC-striatum FC and cortico-amygdalar FC (22; 23). A population-based investigation then showed similar patterns of findings in that FC within emotional regulation network negatively scaled with increasing inflammation composite score (aggregate of CRP, IL-6, IL-10 and TNFα) and classical monocytes (57).

In addition to CRP and IL-6, our investigation on cellular markers and FC showed further evidence for innate immune system involvement as the inflamed-MDD subgroup had increased leukocytes of myeloid origin, especially neutrophils. In particular, we noted negative scaling we observed between neutrophils and FC. Nonetheless, it would be biased and perhaps premature to assert at this stage that only the innate immune system is central to the etiology of inflammation-linked depression. Whilst neutrophils are widely accepted as primary effectors of the innate immune system and acute inflammation, these cells are not functionally exclusive to the innate immune system. Neutrophils engage heavily in immune cross-talk with resident cells of the adaptive immune system, particularly CD4+ T-cells, where mutual modulation of function occurs (33). For example, neutrophils are able to induce activation and promote differentiation of näıve CD4+ T-cells. Reciprocally, regulatory T-cells produce cytokines promoting survival of neutrophils, that otherwise undergo frequent spontaneous apoptosis to facilitate normal cell turnover (58; 59).

In the context of inflammation-linked depression, our evidence suggest that the brain-immune relationship involves interoceptive immune-sensing or simply put, perception of immune state by the brain. This proposition is consistent with evidence supporting the “immunological homunculus” (60; 61; 62) – the concept that discrete neural networks coordinate components of the peripheral immune system via the cholinergic anti-inflammatory pathway, and rostro-caudal functional topography exhibited by the insula. In the human interoceptive model, the posterior, middle and anterior parts of the insular cortex are thought to play different roles. Lower-level (peripheral) sensory information is firstly encoded in the posterior insula, before being represented in the mid-insula where convergent signals, e.g. hedonic/motivational signals to ascribe salience, are integrated with other sensory brain regions. This information is then relayed to the anterior insula, where in conjunction with cingulate cortex, behavioural responses and emotional changes are elicited (48; 63; 64)(48, 63, 64). Thus the posterior insula may be viewed as the immune sensory cortex, whereas the anterior insula is linked to higher-order emotion regulation, such as subjective feeling states of “sickness” and/or “sadness”, and related depressive states such as anhedonia (65; 66) (65, 66). Hence, we interpreted our observations as evidence of dysconnectivity within the INS associated with impairment of immune-sensing and effects on mood regulation in depression.

### Strengths, limitations and conclusions

A strength of our study was the use of NBS for whole-brain analysis FC abnormalities in inflammation-linked depression. Comparable prior studies have generally used a priori defined brain regions and canonical functional networks to ascertain FC alterations. In contrast, NBS allowed us to perform an unbiased wholebrain investigation beyond the constraints of canonical functional networks, which is a more robust approach to mapping effects of inflammation in depression, as networks, clusters and circuits within the brain are more likely affected than single isolated connections. Additionally, we used the Glasser parcellation which is currently the highest definition parcellation scheme for the insular-opercular region, delineating 13 insular/frontal-opercular subdivisions on the basis of a combination of features derived from multiple imaging modalities (67). Our study also presents FC associations with immune markers beyond CRP and IL-6, that is immune cellular markers.

An important limitation of our study is the lack of high CRP (> 3mg/L) controls. An interesting analysis would be to determine if the interoceptive network identified was similarly affected by peripheral inflammation in healthy population and/or abnormal in other recognized interoceptive disorders and immune disorders. It is also important to clarify that although we interpreted our results in relation to prior knowledge of interoceptive systems, we are not asserting that the brain regions identified by the NBS analysis of hiCRP case-control differences are linked exclusively to interoception. Several studies have highlighted the anterior insula as part of a “multiple-demand” system or network (68; 69; 70) and, together with the anterior cingulate and frontal cortex, the insula/operculum has been posited to form a “core” task-dependent brain network responsible for encoding error signals and sustaining attentional control (69). Therefore, more work is needed to strengthen our claim regarding the sensitivity of interoceptive networks to peripheral inflammatory signals, as suggested by these data.

Next, although the sample size was consistent with many prior studies, it was somewhat underpowered to detect the subtle associations between FC and peripheral inflammation. Sampling bias was evident in terms of a greater proportion of females, particularly in the hiCRP MDD cases, although the cases (overall) and controls were prospectively matched for age and sex. In sensitivity analyses, we observed a more circumscribed NBS network **(Figure S5)** with greater effect size when sex was controlled for during NBS testing; and all significant relationships between FC and immune biomarkers within the NBS-defined mask were conserved, albeit with smaller effect size, when sex was statistically controlled in the analysis **(Figure S6)**. In relation to clinical confounds, there was an appreciable effect of BMI in our investigation, indicated by the effects of BMI when included as a covariate in both part 1 and part 2 analyses **(Table S3B-C)**. Finally, we also note that our observations were based on cross-sectional investigation (and not longitudinal), limiting causal interpretation at present.

While the relevance of interoception to inflammation-linked depression has been previously discussed, this study provides direct evidence for brain functional abnormalities in an interoceptive network identified by whole-brain analysis in a clinical MDD sample. These results point towards a putative etiological model of inflammation-linked depression, where peripheral inflammation and depression are linked to dysconnectivity of a brain network specialised in peripheral immune sensing and emotion regulation.

## Data Availability

Further information and requests for resources should be directed to Athina Aruldass. In compliance with Wellcome Open Research, our data will be made available on a scientifically accredited platform upon publication of peer-reviewed final version of the article.

## Acknowledgements

The authors thank all the participants in the study and members of the NIMA research team – in particular, project coordinators Linda Pointon, Junaid Bhatti, Ciara O’Donnell (see Supplementary Appendix for a complete list of NIMA Consortium members), medical imaging staff and laboratory staff. František Vàša provided valuable feedback on an earlier version of the manuscript.

