## Supplemental Data (SD) for "Dysconnectivity of a brain functional network was associated with blood inflammatory markers in depression"

#### *Supplementary Methods*


| | | |
|---|---|---|
| **Section 1** | **Participants** | |
| | **Figure S1** Flow diagram outlining progression of participants through to analyses | **2** |
| **Section 2** | **Blood immune biomarkers** | |
| | Immune biomarker measurement and phenotyping | **3** |
| | **Figure S2 Quality control flow-chart for inflammatory proteins** | **4** |
| | **Figure S3** Data origin flow-chart for Cellular biomarker analyses | **5** |
| **Section 3** | **Neuroimaging** | |
| | **Figure S4** Functional connectivity matrix construction and methodological overview | **6** |
| **Section 4** | **Network-based statistics (NBS)** | |
| | Additional methods | **7** |


#### *Supplementary Results*


| | | |
|---|---|---|
| **Section 5** | **Sample characteristics** | |
| | **Table S1A** Immune biomarker (soluble inflammatory proteins) variation in MDD cases (N=72 MDD) | **8** |
| | **Table S1B** Immune biomarker (cell-counts) variation in MDD cases (N=36 MDD) | **9** |
| | **Table S1C** Cellular biomarker variation in “uninflamed” and “inflamed” MDD subgroup | **10** |
| **Section 6** | **Case-control differences in FC** | |
| | **Table S2A** Constituent edges within HC vs hiCRP NBS network | **11** |
| | **Table S2B** Constituent nodes within HC vs hiCRP NBS network | **12** |
| | **Table S2C** BrainMap database search results (8 pooled studies) | **13** |
| | **Figure S5** Sex adjusted HC vs hiCRP MDD NBS testing | **14** |
| **Section 7** | **Association between FC and immune biomarkers** | |
| | **Table S3A** Edge-wise correlation between FC (of HC vs hiCRP NBS network) and inflammatory proteins | **15** |
| | **Table S3B** Results of hierarchical regression on average network connectivity ~ CRP | **16** |
| | **Table S3C** Results of hierarchical regression on average network connectivity ~ IL-6 and Neutrophils | |
| | **Figure S6A** Sensitivity analyses for NBS testing | **16** |
| | **Figure S7** Association between FC and cellular markers | **17** |

#### Supplemental references **18**

### *Supplemental Methods*

#### Section S1 Participants

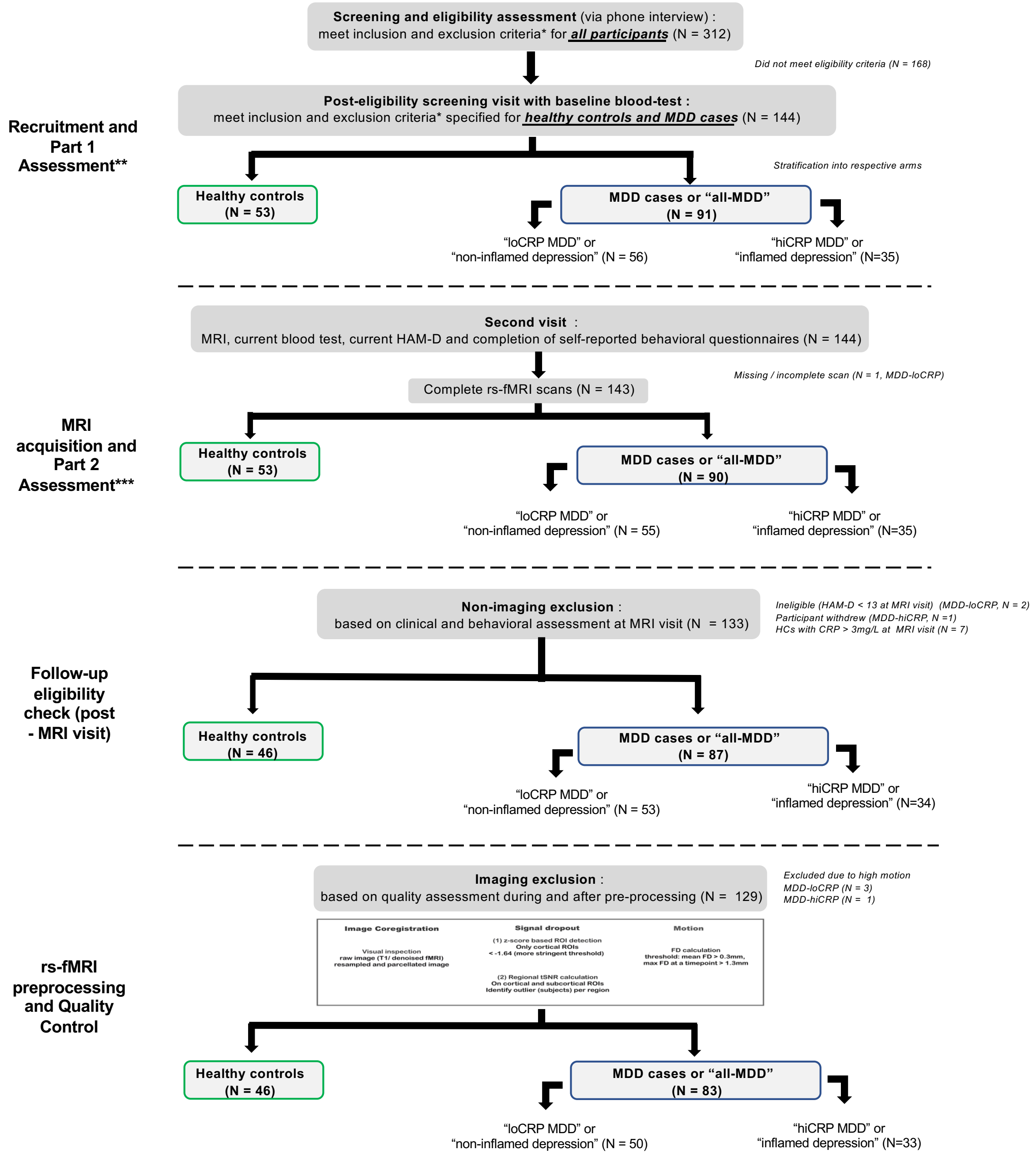


**Figure S1 Flow diagram outlining progression of participants through to analyses.**

### Section S2 Blood immune biomarkers

***Serum and plasma collection*** We collected up to 90mL of venous blood by antecubital venepuncture from participants who had fasted overnight from midnight to between 08:00 and 10:30am on the day of assessment. Participants had refrained from exercise for 72 hours to blood sampling. Blood (8.5ml) were collected into BD Vacutainer® Serum Separator Tubes (SST™ II Advance) Silica tubes and Plasma Preparation Tubes (PPT™) and incubated for 30 minutes at room temperature to allow blood to coagulate. Tubes were centrifuged at 1600 Relative Centrifugal Force (rcf) for 15 minutes and serum or plasma collected in aliquots. All serum samples were stored at -70°C until analysis, with no freeze-thaw cycles.

***hsCRP assay*** Collected blood samples were allowed to coagulate for 30-60 minutes then centrifuged at 1600rcf for 15 mins. 1ml of the resultant sample was then transferred to a white-topped serum tube using a pipette, and transported at room temperature to central laboratory ($Q^2$ solutions). Samples were exposed to anti- CRP-antibodies on latex particles, and the increase in light absorption due to complex formation was used to quantify CRP levels, using Turbidimetry on Beckman Coulter AU analyzers. Both initial hsCRP assay (Part 1 assessment, ***see SA2***) and Part 2 hsCRP were performed at a single central laboratory ($Q^2$ Solutions, The Alba Campus, Livingston EH54 7EG, UK) from 0.5mL of plasma.

***Multiplex cytokine immunoassay*** Cytokine/chemokine levels were measured in plasma and serum using the Chemokine panel 1 (human), Cytokine panel 1 (human) and Pro-inflammatory panel 1 (human) V-PLEX 10-spot immunoassay kits from Meso Scale Discovery (MSD). Samples were assayed in duplicate per manufacturer's instructions. For some cytokines, e.g., IL-1β, no participants had serum or plasma concentrations above the assay's lower limit of detectability (LLOD). Many cytokines also displayed a high inter-assay coefficient of variation (CV) across duplicate measurements (plate-to-plate consistency). Therefore, after first-pass QC on assayed cytokines, we had usable data on 18 cytokines and chemokines collectively (of 30 assayed). Further QC was performed on subset of this data i.e. only MDD cases to detect (i) cytokines with concentrations below assay's lower limit of quantifiability (LLOQ), (ii) intra-assay CV > 30% (identical sample, duplicate runs), and (iii) extreme outliers i.e. concentrations beyond 3 times interquartile range (IQR) per biomarker (**Figure S2**). After final QC, we had analysable data on N=72 MDD cases over 16 cytokines, including IL-6 (**Figure S2**). All values were log-transformed (base 10) to achieve normality for subsequent statistical modelling. For this study, only IL-6 was studied alongside CRP as part of our primary immune variable following established biological associations between the two immune markers (also noted in our dataset, **Table S1A-B**).

***Peripheral blood mononuclear and polymorphonuclear cells immunophenotyping*** Peripheral blood measurements and immunophenotyping have been described in greater detail in a previous study (1). Briefly, flow cytometry was performed on fresh PBMC. Data were manually gated, blind to status of each participant i.e. case/control. We had analysable data on N=36 MDD cases, stratified further into uninflamed and inflamed -MDD subgroups based on clustering analyses described in a preceding study (1) (**Figure S3**, **Table S1C**). We also also used first principal component (PC1) score from the identical study. Principal component analysis was performed over 14 blood cells (12 leukocytes, platelets and red blood cells). A weighted average of cellular variables, PC1 accounted for 19% of total variance and was more strongly weighted on cells of myeloid origin, namely neutrophils, basophils and classical monocytes.

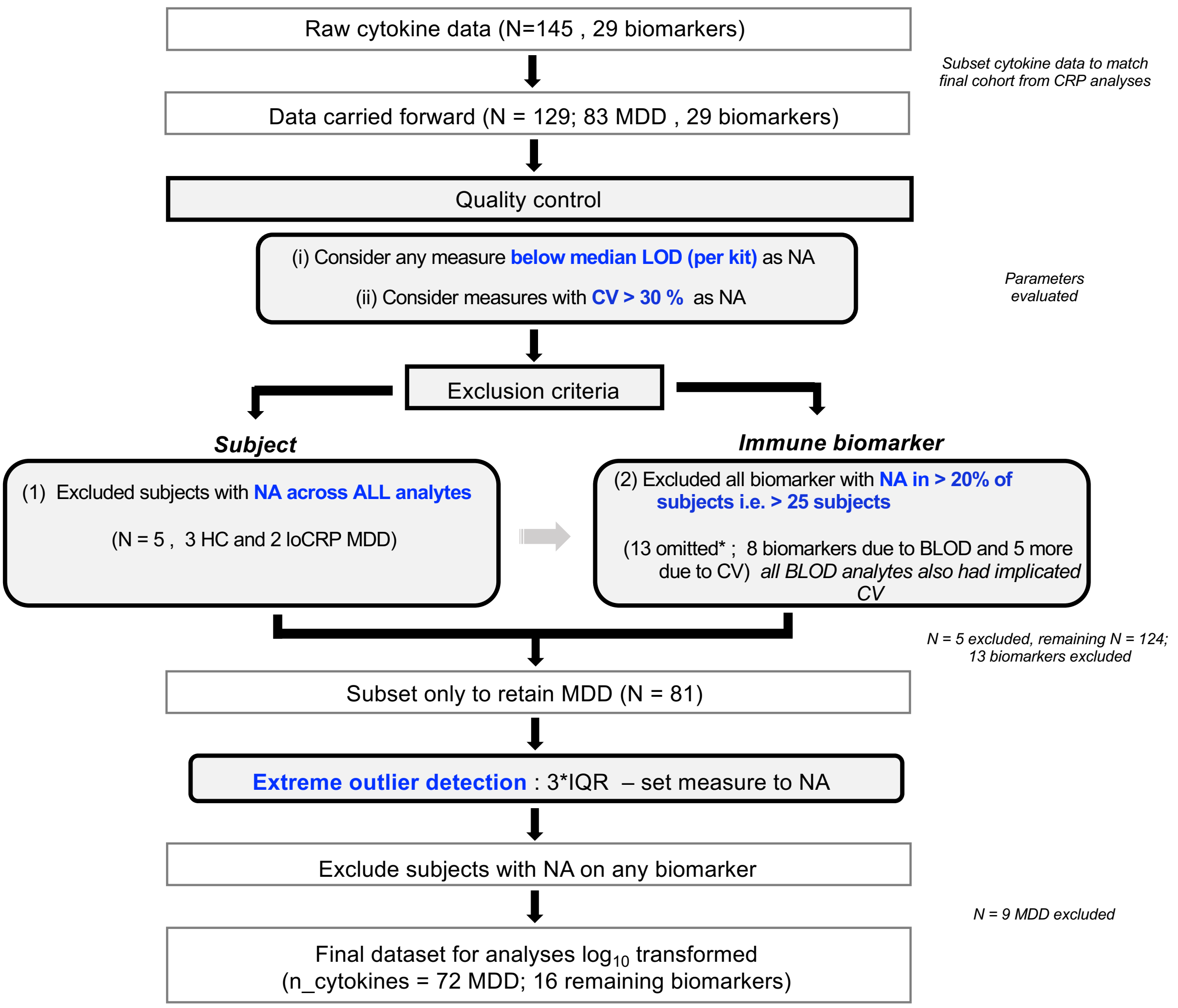


**Figure S2 Quality control flow-chart for inflammatory proteins (N=72 MDD).**

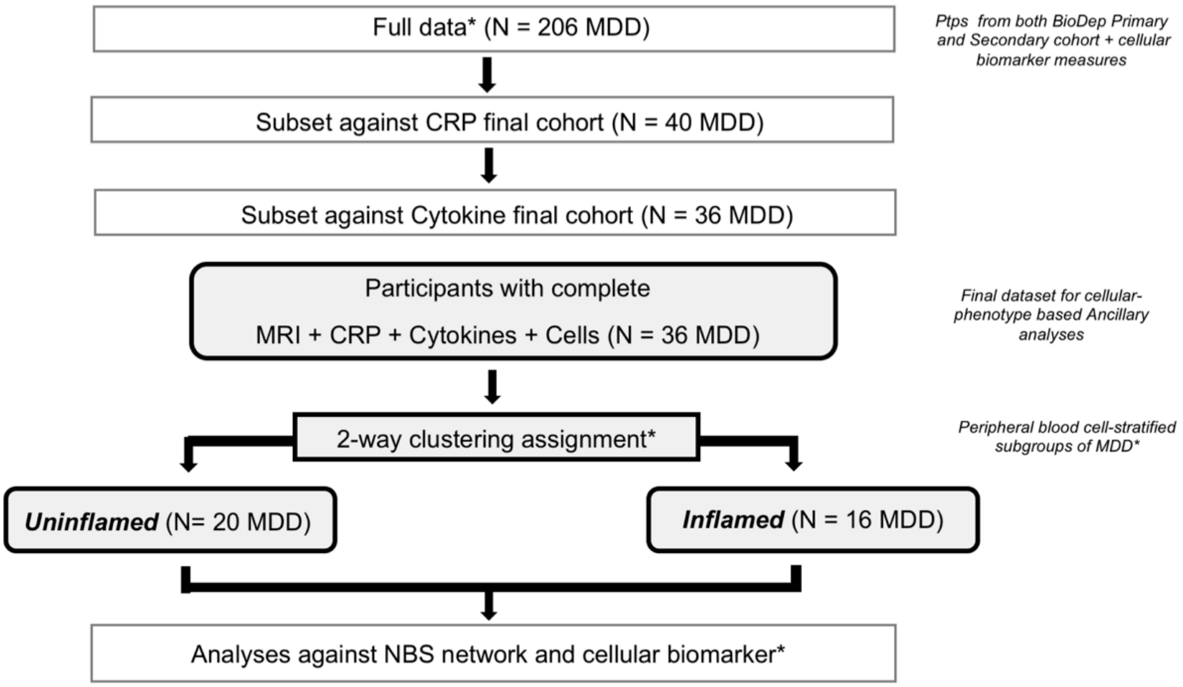


**Figure S3 Data origin flow-chart for cellular biomarker analyses (N=36 MDD).** *from previous study by Lynall et al. (2020). Cell-counts were available across 12 peripheral blood mononuclear and polymorphonuclear cells.

***Electronic Data Management*** Study data e.g. sociodemographic information, immune markers, were collected and managed for central access using REDCap (Research Electronic Data Capture) electronic data capture platform hosted at the University of Cambridge (2).

### Section S3 Neuroimaging

A

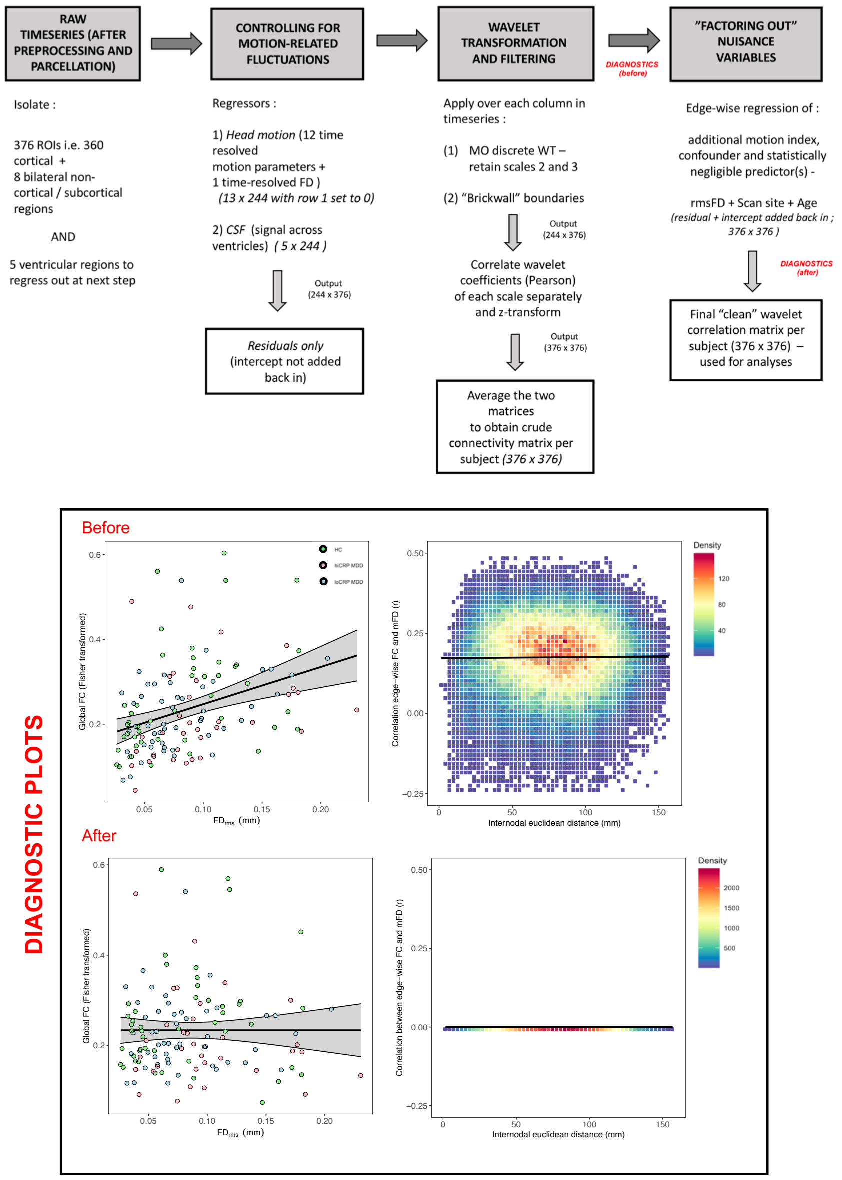


B

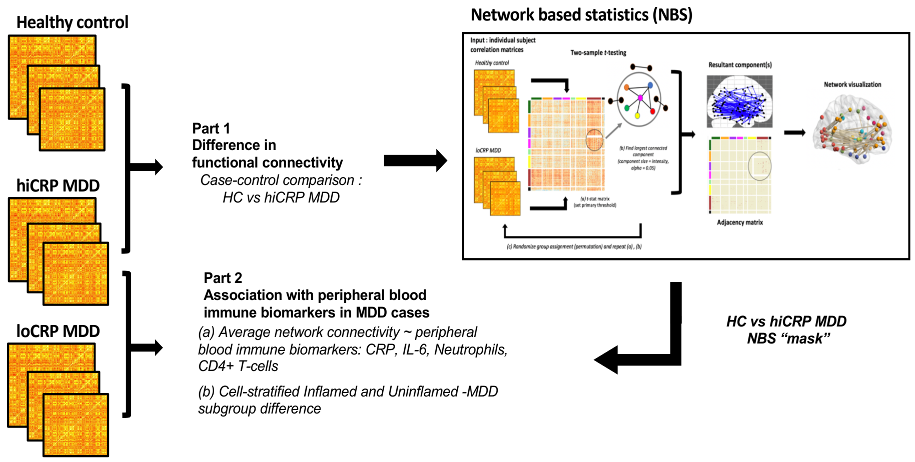

**Figure S4 Functional connectivity matrix construction and methodological overview. (A)** After standard preprocessing of fMRI timeseries, we critically took additional measures to ascertain any residual motion-related effects on functional connectome. Our diagnostic procedure comprised (i) assessing for effects of motion (indexed by root mean square framewise displacement; $FD_{rms}$) on global FC, and (ii) determining likelihood of edge-wise correlation between FC and mean FD, and if this association fluctuated with internodal distance† i.e. distance between two connecting nodes, as previously demonstrated (3–5). From the diagnostic plots (*Before*) we noted (i) a global effect of motion on FC, in that global FC increased with motion ($\beta$ = 0.88, $R^2_{adj}$ = 0.13, $p$ = 2x10$^{-5}$), and (ii) on average edges showed correlation with mean FD (indicated by *y*-intercept = 0.17) and this correlation increased marginally (positive slope; $\beta$ = 0.00004) with increasing distance between nodes i.e. long-range edges were more vulnerable to motion-related effects compared to short-range edges ($R^2_{adj}$ = 8.11x10$^{-5}$, $p$ = 0.01). After edge-wise regression of $FD_{rms}$ (*After*), we remedied the previously observed (i) global relationship between FC and motion ($\beta$ = 0.0, $R^2_{adj}$ = -0.01, $p$ = 1.0), and (ii) edge-level correlation between FC and motion (*y*-intercept = 7.9x10$^{-18}$) and fluctuation due to internodal distance ($\beta$ = -1.38x10$^{-20}$, $R^2_{adj}$ = 8.87x10$^{-5}$, $p$ = 0.007). **(B)** Two-part analytic pipeline. †distance spanning across nodes was defined as Euclidean distance; $C_{[i,j]} = \sqrt{((\Delta X)^2 + (\Delta Y)^2 + (\Delta Z)^2)}$; where $\Delta X = X_i - X_j$ , *i* = node 1, *j* = node 2; X,Y and Z = centroid coordinates in MNI space for each mapped parcel.**p* < .05; ***p* < .01; ****p* < .001

### Section S4 Network-based statistics (NBS)

Network-based statistics (NBS) (7) was implemented in the NBS Toolbox for Matlab R2017. The method was designed to circumvent the multiple-comparisons problem associated with mass univariate testing on connectivity networks, for example at nodal and individual edge-level. NBS instead performs a single global statistical test on subsets of connected nodes i.e. “cluster(s)” or subnetwork(s). Therefore, whilst NBS perhaps bypasses the statistical problem accompanying connectivity analyses, it does so at the cost of anatomical precision. Since resultant p-value is ascribed to the emergent subnetwork, experimental inferences can pertain only to the component and not the constituent edges. Hypothesis testing with NBS is a 2-step process broadly involving (i) edge-wise general linear model fitting for test-statistic estimation e.g. t-statistic estimation with one-tailed t-testing, and (ii) permutation testing. Briefly, a primary component-forming threshold i.e. t-statistic threshold (at p <.05, uncorrected) is applied to identify edges displaying differences in functional connectivity strength. Edges exceeding this test threshold or supra-threshold connections form a “cluster” in topological space, modelling the uncorrected observed experimental effects. At this stage also, the maximum component size is defined either using extant (number of edges) or intensity (sum of t-statistic i.e. approximate effect size). Permutation testing is then applied repeating the above process, with group assignments on connectivity matrices permuted each time. Maximum component size is computed each time to build sample distribution. Following permutation testing, the global statistical test is carried out to determine statistical significance of initially observed component. This is defined as the ratio of permutations yielding a component with equal or greater size than observed component, correcting for the family-wise error rate at cluster level with p < .05 (see equation below).

$$p - value = \frac{number\ of\ permutations\ with\ maximum\ component\ size\ \geq observed\ component\ size}{number\ of\ permutations}$$

### *Supplemental Results*

#### Section S5 Sample characteristics

| Variable | Mean (s.d.) (N=72) | Correlation with CRP (Pearson's r) | p-value (FDR adj.) |
|---|---|---|---|
| Sex, Male (n, %) | 22 (30.6) | - | - |
| BMI ($kg/m^2$) | 26.74 (4.05) | 0.48 | $<$**0.001***** |
| **Inflammatory markers** | | | |
| CRP ($log_{10}$ $mg/L$) | 0.21 (0.51) | - | - |
| IL-6 ($log_{10}$ $pg/L$) | -0.22 (0.27) | 0.62 | $<$**0.001***** |

**Table S1A Immune biomarker (soluble inflammatory proteins) variation in MDD cohort (N=72 MDD).** Eleven MDD participants (N=11 MDD) were excluded from initial analyzable data i.e. final cohort for functional connectivity and CRP analyses, following QC of cytokine data. For purpose of the present investigation, only IL-6 was analysed further. FDR adj.; false discovery rate adjusted; IL; interleukin; .**p* < .05; ***p* < .01; ****p* < .001

| | Mean (N=36) | SD | Correlation with immune biomarkers | | | |
|---|---|---|---|---|---|---|
| | | | CRP | | IL-6 | |
| | | | *r* | $p_{FDR}$ | *r* | $p_{FDR}$ |
| BMI (kg/m$^2$) | 27.05 | 4.01 | 0.57 | **0.001** | 0.45 | $<$ **0.05*** |
| **Inflammatory proteins** | | | | | | |
| CRP[a] | 0.2 | 0.49 | - | - | 0.64 | $<$ **0.001***** |
| IL-6[b] | -0.26 | 0.29 | 0.64 | $<$ **0.001***** | - | - |
| **Cellular biomarkers (x10$^3$ $\mu$/L)** | | | | | | |
| Basophils | 0.03 | 0.02 | 0.16 | 0.60 | -0.01 | 1 |
| Eosinophils | 0.2 | 0.2 | 0.35 | 0.13 | 0 | 1 |
| Neutrophils | 3.9 | 1.46 | 0.57 | $<$ **0.001***** | 0.47 | $<$ **0.05*** |
| Classical monocytes | 0.39 | 0.17 | 0.14 | 0.60 | 0.16 | 0.66 |
| Nonclassical monocytes | 0.05 | 0.03 | 0.1 | 0.70 | 0.06 | 1 |
| Intermediate monocytes | 0.01 | 0.02 | 0.16 | 0.60 | 0.14 | 0.72 |
| CD4+ T-cells | 1.06 | 0.34 | 0.27 | 0.33 | 0.25 | 0.35 |
| CD8+ T-cells | 0.43 | 0.22 | 0.01 | 1 | 0.27 | 0.29 |
| B-cells | 0.09 | 0.08 | 0.1 | 0.70 | 0.22 | 0.42 |
| CD16+ NK cells | 0.1 | 0.09 | 0.14 | 0.60 | 0.12 | 0.74 |
| CD56+ NK cells | 0.01 | 0.01 | 0.25 | 0.33 | 0.32 | 0.17 |
| NK T-cells | 0.08 | 0.09 | 0.15 | 0.60 | -0.02 | 1 |

**Table S1B Correlation between cell-counts and primary immune biomarkers in MDD cases prior to "uninflamed" and "inflamed" subgroup stratification (N=36 MDD).**
[a]log10 mg/L; [b]log10 pg/L; $P_{FDR}$; false discovery rate adjusted p-value; *p < .05; **p < .01; ***p < .001 .

| | **Uninflamed** (N=20) | | **Inflamed** (N=16) | | **p-value**[a] |
|---|---|---|---|---|---|
| | Mean | SD | Mean | SD | |
| **Sociodemographic / Clinical** | | | | | |
| Age (years) | 36.46 | 7.75 | 38 | 6.48 | 0.52 |
| BMI (kg/$m^2$) | 25.94 | 3.52 | 28.43 | 4.25 | 0.07 |
| Sex, Male (n, %) | 7 | 35 | 4 | 25 | 0.52 |
| CRP[b] | 0.00 | 0.46 | 0.44 | 0.43 | $<$ **0.01**** |
| IL-6[b] | -0.34 | 0.24 | -0.16 | 0.32 | 0.07 |
| **Cellular biomarkers (x$10^3$ $\mu$/L)** | | | | | |
| Basophils | 0.02 | 0.02 | 0.05 | 0.03 | $<$ **0.01**** |
| Eosinophils | 0.12 | 0.08 | 0.31 | 0.26 | **0.01**** |
| Neutrophils | 3.12 | 1.01 | 4.87 | 1.35 | $<$ **0.001***** |
| Classical monocytes | 0.32 | 0.1 | 0.47 | 0.2 | $<$ **0.01**** |
| Nonclassical monocytes | 0.04 | 0.02 | 0.06 | 0.04 | 0.16 |
| Intermediate monocytes | 0.01 | 0.01 | 0.02 | 0.03 | 0.13 |
| CD4+ T-cells | 1.06 | 0.32 | 1.07 | 0.37 | 0.88 |
| CD8+ T-cells | 0.41 | 0.14 | 0.46 | 0.3 | 0.58 |
| B-cells | 0.08 | 0.06 | 0.1 | 0.1 | 0.65 |
| CD16+ NK cells | 0.08 | 0.04 | 0.13 | 0.12 | 0.17 |
| CD56+ NK cells | 0.01 | 0.01 | 0.02 | 0.01 | 0.54 |
| NK T-cells | 0.05 | 0.03 | 0.12 | 0.13 | $<$ **0.05*** |

**Table S1C Cell-count variation in MDD cases (N=36 MDD total).** Depending on full biomarker measurement availability i.e. CRP, inflammatory proteins, cell-count, N=36 MDD cases were stratified further into "Uninflamed" and "Inflamed" subgroups based on clustering analyses outcome reported in Lynall et al. (2020). [a]uncorrected p-value from Welch's t-test i.e. not corrected for multiple cell-count comparison; [b]$\log_{10}$ transformed; *p < .05; **p < .01; ***p < .001 .

### Section S6 Case-control differences in FC

| Edge | Node pair [i , j] | | Func. mod.[a] [i , j] | Anat. reg.[b] [i , j] | t–stat |
|---|---|---|---|---|---|
| # | Node_i | Node_j | Yeo7-cort[c] / FS_Subcort[d] | Glasser_Cort[e] / FS_Subcort | |
| 1 | L_d23ab | L_FOP4 | DMN – VA | PCC – Ins_fOperc | 4.423 |
| 2 | L_31pv | L_FOP4 | DMN – VA | PCC – Ins_fOperc | 4.304 |
| 3 | R_8Ad | L_FOP1 | DMN – VA | dlPFC – postOperc | 4.238 |
| 4 | R_PGs | R_33pr | DMN – VA | infPar – ACC_mPFC | 4.017 |
| 5 | R_d23ab | R_33pr | DMN – VA | PCC – ACC_mPFC | 3.996 |
| 6 | L_POS1 | L_FOP4 | DMN – VA | PCC – Ins_fOperc | 3.991 |
| 7 | R_8Ad | L_FOP4 | DMN – VA | dlPFC – Ins_fOperc | 3.926 |
| 8 | L_STSvp | L_FOP4 | DMN – VA | dlPFC – Ins_fOperc | 3.849 |
| 9 | R_RSC | L_PF | DMN – VA | PCC – Ins_fOperc | 3.838 |
| 10 | L_POS1 | L_FOP1 | DMN – VA | PCC – postOperc | 3.822 |
| 11 | R_d23ab | R_A1 | DMN – SM | PCC – EarlyAud | 4.19 |
| 12 | R_d23ab | R_OP4 | DMN – SM | PCC – postOperc | 4.078 |
| 13 | R_d23ab | L_LBelt | DMN – SM | PCC – EarlyAud | 4.059 |
| 14 | L_d23ab | R_A1 | DMN – SM | PCC – EarlyAud | 4.019 |
| 15 | L_31pv | R_A1 | DMN – SM | PCC – EarlyAud | 4.004 |
| 16 | R_d23ab | L_MBelt | DMN – SM | PCC – EarlyAud | 3.887 |
| 17 | L_d23ab | R_OP4 | DMN – SM | PCC – postOperc | 3.872 |
| 18 | R_d23ab | L_PBelt | DMN – SM | PCC – EarlyAud | 3.813 |
| 19 | R_IP1 | R_33pr | FP – VA | infPar – ACC_mPFC | 4.27 |
| 20 | L_7Pm | L_MI | FP – VA | supPar – Ins_fOperc | 4.037 |
| 21 | L_7Pm | L_FOP1 | FP – VA | supPar – postOperc | 3.899 |
| 22 | L_IP2 | L_MI | FP – VA | infPar – Ins_fOperc | 3.861 |
| 23 | L_IP2 | L_6r | FP – VA | infPar – Premotor | 3.844 |
| 24 | R_Thalamus | L_PFm | Subcort – FP | Subcort – infPar | 4.139 |
| 25 | R_Thalamus | L_IP2 | Subcort – FP | Subcort – infPar | 4.011 |
| 26 | L_Thalamus | L_7Pm | Subcort – FP | Subcort – supPar | 3.998 |
| 27 | L_Thalamus | L_IP2 | Subcort – FP | Subcort – infPar | 3.917 |
| 28 | L_Thalamus | L_PFm | Subcort – FP | Subcort – infPar | 3.808 |
| 29 | L_s32 | L_MT | DMN – Visual | ACC_mPFC – MTcompNeighVis | 3.859 |
| 30 | L_31pv | L_LO3 | DMN – Visual | PCC – MTcompNeighVis | 3.901 |
| 31 | L_s32 | L_LO3 | DMN – Visual | ACC_mPFC – MTcompNeighVis | 4.381 |
| 32 | R_PGs | R_H | DMN – Subcort | infPar – medTemp | 3.984 |
| 33 | L_POS1 | L_Putamen | DMN – Subcort | PCC – Subcort | 3.931 |
| 34 | R_8Ad | L_Putamen | DMN – Subcort | dlPFC – Subcort | 3.876 |
| 35 | L_6r | L_7PL | VA – DA | Premotor – postOperc | 4.478 |
| 36 | R_Thalamus | L_PF | Subcort – VA | Subcort – Ins_fOperc | 4.109 |
| 37 | L_PFm | L_PGp | FP – DA | infPar – Ins_fOperc | 3.95 |
| 38 | L_s32 | L_9a | DMN – DMN | ACC_mPFC – dlPFC | 3.88 |

**Table S2A Constituent edges within HC-hiCRP NBS network.** [a]Functional module; [b]Anatomical region; [c]Yeo-7 cortical functional network as defined by Yeo et al. (2011) (8); [d]*FreeSurfer* derived subcortical segments (9); [e]Glasser et al. (2016) 22 cortical regions (10). Abbreviations: DMN – default mode network; VA – ventral attentional network; SM – somatosensory and somatomotor network; DA – dorsal attentional network; FP – fronto-parietal control; PCC – posterior cingulate cortex; postOperc – posterior opercular cortex; dlPFC – dorsolateral prefrontal cortex; Ins_fOperc – insular and frontal opercular cortex; infPar – inferior parietal cortex; ACC_mPFC – anterior cingulate and medial prefrontal cortex; medTemp – medial temporal cortex; supPar – superior parietal cortex; EarlyAud – early auditory cortex; MTcompNeighVis – middle temporal+ complex and neighbouring visual area; Premotor – premotor cortex. Edges shown in Figure S5A(I), S5A(II).

| # | ROI label[a] | Centroid coordinates[b] | | |
|---|---|---|---|---|
| | | x | y | z |
| 1 | Left-Thalamus-Proper | -11.75 | -19.88 | 5.17 |
| 2 | Left-Putamen | -26.37 | -0.16 | -2.09 |
| 3 | Right-Thalamus-Proper | 12.49 | -19.23 | 5.66 |
| 4 | L_MT | -45.44 | -73.11 | 6.03 |
| 5 | L_7Pm | -6.63 | -66.01 | 48.22 |
| 6 | L_POS1 | -10.66 | -59.16 | 12.68 |
| 7 | L_d23ab | -3.20 | -40.42 | 30.45 |
| 8 | L_31pv | -8.38 | -45.21 | 32.26 |
| 9 | L_7PL | -12.25 | -71.86 | 51.38 |
| 10 | L_6r | -52.10 | 5.69 | 17.11 |
| 11 | L_9a | -19.30 | 53.80 | 24.56 |
| 12 | L_FOP4 | -40.14 | 11.90 | 6.10 |
| 13 | L_MI | -36.12 | 9.04 | 1.20 |
| 14 | L_FOP1 | -50.02 | 2.60 | 3.78 |
| 15 | L_PBelt | -54.11 | -24.86 | 5.04 |
| 16 | L_STSvp | -55.27 | -35.68 | -6.72 |
| 17 | L_PGp | -40.21 | -81.15 | 21.38 |
| 18 | L_IP2 | -39.54 | -51.38 | 38.24 |
| 19 | L_PF | -56.89 | -38.88 | 35.64 |
| 20 | L_PFm | -48.11 | -57.78 | 39.30 |
| 21 | L_LO3 | -45.63 | -79.08 | 8.78 |
| 22 | L_s32 | -6.88 | 34.17 | -13.77 |
| 23 | L_MBelt | -45.12 | -17.79 | 1.67 |
| 24 | L_LBelt | -45.93 | -27.19 | 6.39 |
| 25 | R_RSC | 6.13 | -37.07 | 19.99 |
| 26 | R_A1 | 44.06 | -23.08 | 9.56 |
| 27 | R_d23ab | 3.55 | -40.79 | 31.40 |
| 28 | R_33pr | 4.18 | 10.74 | 29.61 |
| 29 | R_8Ad | 22.78 | 27.38 | 43.91 |
| 30 | R_OP4 | 58.35 | -13.61 | 15.65 |
| 31 | R_H | 31.62 | -31.45 | -11.51 |
| 32 | R_IP1 | 34.65 | -65.75 | 42.25 |
| 33 | R_PGs | 42.88 | -67.22 | 38.01 |

**Table 2B Constituent nodes within HC-hiCRP MDD NBS network.** [a]per Glasser *et al.* (2016) parcellation scheme and FreeSurfer subcortical segmentation, [b]MNI coordinates.

| Author (year) | Context | Type of fMRI study & task / condition | Sample | Conclusions | Brain regions |
|---|---|---|---|---|---|
| Salomons et al. (2004) | Subjective pain perception and emotional awareness | Task fMRI – pain monitoring | Healthy individuals | Pain perceived as controllable resulted in attenuation in brain areas linked with pain processing. | anterior cingulate, insula, secondary somatosensory cortices |
| de Araujo et al. (2004) | Subjective gustatory perception | Task fMRI – taste, rating oral stimuli for thickness, fat content and sweetness after swallowing | Healthy individuals | Perception of oral stimuli i.e. oral sensory representation of food is represented in the brain (activation of brain regions) and may play a role in hedonic responses to food. | anterior insula, midinsula, dorsal midanterior cingulate cortex and ventral anterior cingular cortex. |
| Botvinick et al. (2005) | Subjective pain perception and emotional awareness | Task fMRI – pain monitoring and viewing facial expressions | Healthy individuals | Common neural pathways and brain regions are engaged in perception of one's own and other's affective states. | Anterior cingulate cortex and insula |
| Henderson et al. (2006) | Subjective pain perception and emotional awareness | Task fMRI - saline injection inducing "deep" and "superficial" pain | Healthy individuals | Distinct sets of brain regions were activated in perception of pain of different intensity and invocation of emotion. | Cingulate cortex, somatosensory cortex, motor cortex |
| Christoffels et al. (2007) | Self-speech monitoring | Task fMRI – overt naming of visual stimuli, followed by auditory feedback of own voice. | Healthy individuals | Auditory feedback processing involves a network of disparate regions related to performance monitoring and speech-motor control. | Cingulate cortex, bilateral insula, supplementary motor area, cerebellum, thalamus, basal ganglia |
| Samson et al. (2011) | External emotional perception and emotional awareness | Task fMRI - passive viewing of visual stimuli i.e. sad faces and houses | Healthy controls and Major depressive disorder (MDD) patients; stratified into treatment responders and non-responders | Treatment success in MDD might be related to optimal self-referential and emotionally salient stimuli processing within the brain. | Posterior cingulate cortex, dorsomedial prefrontal cortex, superior frontal gyrus, caudate nucleus and insula |
| Johnstone et al. (2012) | Subjective pain perception and emotional awareness | Task fMRI – detection of thermal tactile stimulus, respond with button press if perceived as painful | Healthy individuals | Activation of neural regions by which nociceptive signals become salient, corresponded with intensity of thermal stimuli and self-reported painful experience. | Insula, mid-cingulate, ventral lateral thalamic nuclei, globus pallidus and premotor cortex |
| Smieskova et al. (2015) | Subjective motivational / reward processing | Task fMRI – Salience Attribution Task was completed; response to high-probability rewarding cues / relevant cues (adaptive salience) and response to low-probability rewarding cues / irrelevant cues (aberrant salience) was measured through brain activity. | Healthy controls and Schizophrenia patients; stratified into at-risk, first-episode psychosis, and nonschizophrenic psychosis | Antipsychotics modulate neural cingulo-insular in motivational salience processing. | Cingulate cortices, insular cortex, secondary somatomotor cortex, premotor cortex |

**Table S2C Output from BrainMap database search for studies matching "insula" and "cingulate" or "cingulate cortex" keyword criteria.**

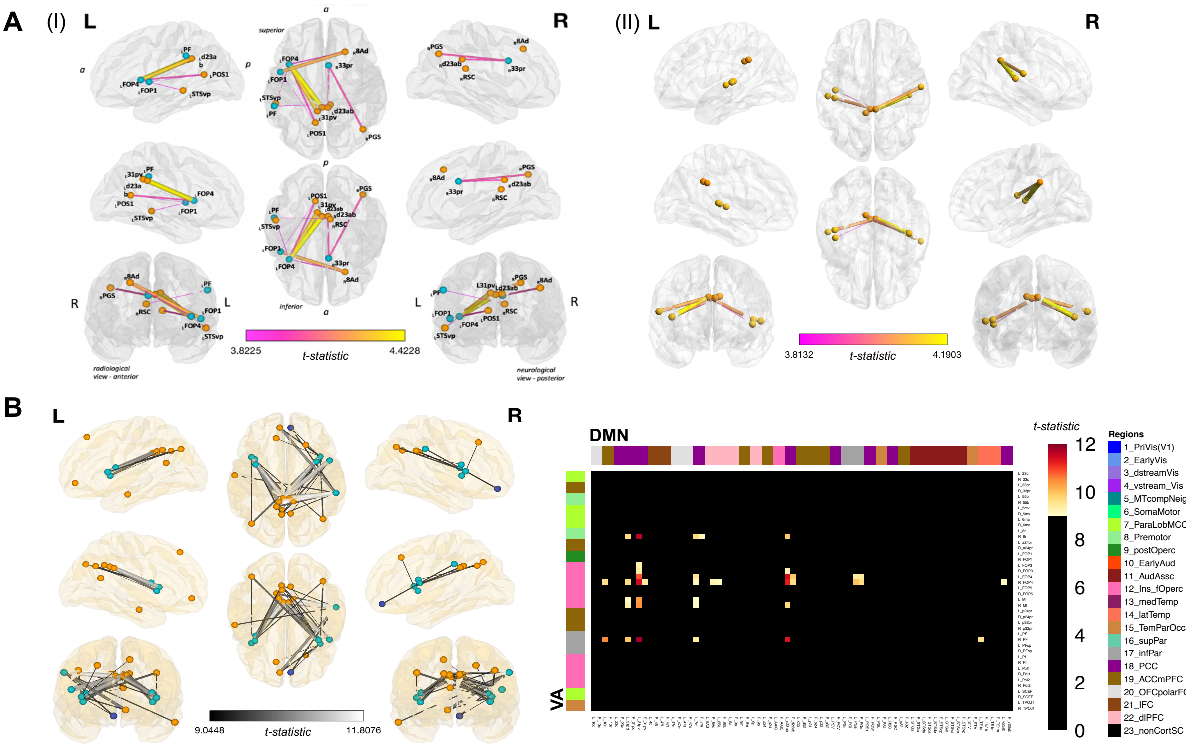


**Figure S5 Network-based case-control differences in FC. (A)** Edges within DMN-VA and DMN-SM shown here. Most edges were lateral-medial, with more localized to the left hemisphere. **(B)** Sex-adjusted HC vs hiCRP MDD NBS network. Constituent edges were almost exclusively located within DMN-VA intermodule, with greater experimental effect (t-stat) range across edges.

### Section 7 Association between functional connectivity and immune biomarkers

| | Node pair for edge | | CRP | | | IL-6 | | | Neutrophils | | |
|---|---|---|---|---|---|---|---|---|---|---|---|
| # | Node_i | Node_j | Corr[a] | p | $p_{FDR}$ | Corr[a] | p | $p_{FDR}$ | Corr[a] | p | $p_{FDR}$ |
| 1 | L_d23ab | L_FOP4 | -0.325 | 0.004 | **0.016** | -0.433 | 0.000 | **0.004** | -0.355 | 0.034 | 0.208 |
| 2 | L_31pv | L_FOP4 | -0.306 | 0.006 | **0.022** | -0.485 | 0.000 | **0.001** | -0.364 | 0.029 | 0.208 |
| 3 | R_8Ad | L_FOP1 | -0.274 | 0.015 | **0.032** | -0.305 | 0.012 | 0.055 | -0.231 | 0.176 | 0.417 |
| 4 | R_PGs | R_33pr | -0.257 | 0.022 | **0.041** | -0.283 | 0.019 | 0.072 | -0.332 | 0.048 | 0.227 |
| 5 | R_d23ab | R_33pr | -0.292 | 0.009 | **0.025** | -0.217 | 0.071 | 0.146 | -0.210 | 0.218 | 0.436 |
| 6 | L_POS1 | L_FOP4 | -0.188 | 0.093 | 0.134 | -0.313 | 0.010 | 0.053 | -0.411 | 0.013 | 0.208 |
| 7 | R_8Ad | L_FOP4 | -0.272 | 0.015 | **0.032** | -0.428 | 0.000 | **0.004** | -0.347 | 0.038 | 0.208 |
| 8 | L_STSvp | L_FOP4 | -0.309 | 0.006 | **0.022** | -0.425 | 0.000 | **0.004** | -0.360 | 0.031 | 0.208 |
| 9 | R_RSC | L_PF | -0.121 | 0.277 | 0.301 | -0.155 | 0.197 | 0.267 | -0.239 | 0.160 | 0.405 |
| 10 | L_POS1 | L_FOP1 | -0.131 | 0.239 | 0.268 | -0.252 | 0.036 | 0.103 | -0.251 | 0.140 | 0.395 |
| 11 | R_d23ab | R_A1 | -0.250 | 0.026 | **0.046** | -0.196 | 0.104 | 0.197 | -0.120 | 0.487 | 0.660 |
| 12 | R_d23ab | R_OP4 | -0.180 | 0.107 | 0.146 | -0.160 | 0.182 | 0.257 | -0.190 | 0.268 | 0.459 |
| 13 | R_d23ab | L_LBelt | -0.273 | 0.015 | **0.032** | -0.216 | 0.073 | 0.146 | -0.166 | 0.333 | 0.527 |
| 14 | L_d23ab | R_A1 | -0.187 | 0.095 | 0.134 | -0.165 | 0.171 | 0.257 | -0.194 | 0.256 | 0.459 |
| 15 | L_31pv | R_A1 | -0.135 | 0.226 | 0.260 | -0.250 | 0.038 | 0.103 | -0.115 | 0.503 | 0.660 |
| 16 | R_d23ab | L_MBelt | -0.372 | 0.001 | **0.007** | -0.287 | 0.017 | 0.072 | -0.134 | 0.435 | 0.636 |
| 17 | L_d23ab | R_OP4 | -0.139 | 0.212 | 0.252 | -0.160 | 0.182 | 0.257 | -0.247 | 0.146 | 0.395 |
| 18 | R_d23ab | L_PBelt | -0.230 | 0.040 | 0.068 | -0.161 | 0.180 | 0.257 | -0.186 | 0.278 | 0.459 |
| 19 | R_IP1 | R_33pr | -0.278 | 0.013 | **0.032** | -0.273 | 0.024 | 0.082 | -0.274 | 0.106 | 0.380 |
| 20 | L_7Pm | L_MI | -0.377 | 0.001 | **0.007** | -0.352 | 0.004 | **0.023** | -0.199 | 0.245 | 0.459 |
| 21 | L_7Pm | L_FOP1 | -0.480 | 0.000 | **0.001** | -0.494 | 0.000 | **0.001** | -0.214 | 0.211 | 0.436 |
| 22 | L_IP2 | L_MI | -0.359 | 0.001 | **0.009** | -0.221 | 0.066 | 0.146 | -0.218 | 0.202 | 0.436 |
| 23 | L_IP2 | L_6r | -0.223 | 0.047 | 0.074 | -0.181 | 0.133 | 0.229 | -0.352 | 0.035 | 0.208 |
| 24 | R_Thalamus | L_PFm | -0.177 | 0.114 | 0.149 | -0.074 | 0.539 | 0.603 | -0.099 | 0.565 | 0.693 |
| 25 | R_Thalamus | L_IP2 | -0.302 | 0.007 | **0.022** | -0.129 | 0.285 | 0.360 | -0.073 | 0.672 | 0.798 |
| 26 | L_Thalamus | L_7Pm | -0.404 | 0.000 | **0.004** | -0.267 | 0.027 | 0.085 | -0.008 | 0.961 | 0.961 |
| 27 | L_Thalamus | L_IP2 | -0.426 | 0.000 | **0.003** | -0.226 | 0.061 | 0.145 | -0.110 | 0.521 | 0.660 |
| 28 | L_Thalamus | L_PFm | -0.299 | 0.008 | **0.023** | -0.184 | 0.126 | 0.229 | -0.047 | 0.786 | 0.854 |
| 29 | L_s32 | L_MT | -0.078 | 0.486 | 0.513 | -0.121 | 0.315 | 0.374 | -0.119 | 0.490 | 0.660 |
| 30 | L_31pv | L_LO3 | -0.022 | 0.843 | 0.843 | -0.011 | 0.925 | 0.925 | 0.054 | 0.756 | 0.845 |
| 31 | L_s32 | L_LO3 | -0.075 | 0.504 | 0.518 | -0.054 | 0.655 | 0.711 | -0.026 | 0.882 | 0.909 |
| 32 | R_PGs | R_H | -0.267 | 0.017 | **0.035** | -0.166 | 0.166 | 0.257 | -0.025 | 0.885 | 0.909 |
| 33 | L_POS1 | L_Putamen | -0.172 | 0.123 | 0.156 | -0.085 | 0.478 | 0.551 | -0.137 | 0.427 | 0.636 |
| 34 | R_8Ad | L_Putamen | -0.207 | 0.065 | 0.099 | -0.125 | 0.298 | 0.365 | -0.269 | 0.112 | 0.380 |
| 35 | L_6r | L_7PL | -0.339 | 0.003 | **0.014** | -0.148 | 0.217 | 0.285 | -0.369 | 0.027 | 0.208 |
| 36 | R_Thalamus | L_PF | -0.333 | 0.003 | **0.015** | -0.033 | 0.780 | 0.824 | -0.264 | 0.120 | 0.380 |
| 37 | L_PFm | L_PGp | -0.226 | 0.044 | 0.072 | -0.230 | 0.056 | 0.141 | -0.294 | 0.082 | 0.347 |
| 38 | L_s32 | L_9a | -0.152 | 0.173 | 0.212 | -0.014 | 0.909 | 0.925 | -0.067 | 0.698 | 0.804 |

**Table S3A Edge-wise correlation between FC (of HC vs hiCRP MDD NBS network) and immune biomarkers.** $p_{FDR} < 0.05$ are in bold. FDR surviving edge common to CRP and IL-6 are highlighted in gray (N=6). 21 edges survived correction for CRP, 6 for IL-6 and none for neutrophils. [a]Fisher transformed Pearson correlation between edge FC and immune biomarker.

| | Model 1 | | Model 2 | | Model 3 | | Model 4 | |
|---|---|---|---|---|---|---|---|---|
| Predictor | $\beta$ | *p* | $\beta$ | *p* | $\beta$ | *p* | $\beta$ | *p* |
| CRP | -0.097 | *** | -0.102 | *** | -0.074 | *** | -0.073 | *** |
| BMI | | | 0.003 | 0.400 | 0.002 | 0.623 | 0.002 | 0.644 |
| Number of antidepressant(s) | | | -0.035 | 0.115 | -0.038 | 0.068 | -0.039 | 0.068 |
| Number of comorbidities | | | -0.024 | 0.322 | -0.027 | 0.240 | -0.027 | 0.245 |
| Number of non-psychotropic drug(s) | | | 0.013 | 0.430 | 0.019 | 0.228 | 0.020 | 0.223 |
| Sex (MALE) | | | | | 0.089 | 0.001** | 0.091 | 0.001** |
| Tobacco (CURRENT) | | | | | 0.021 | 0.512 | 0.021 | 0.518 |
| Alcohol (CURRENT) | | | | | -0.022 | 0.388 | -0.022 | 0.392 |
| Severity(HAMD-17 items) | | | | | | | -0.001 | 0.782 |

**Table S3B Results of hierarchical regression on average network connectivity (derived from HC vs hiCRP MDD NBS network) against CRP.** [a]$\log_{10}$ mg/L; [b]$\log_{10}$ pg/L; [c]body mass index, kg/m$^2$; **p* < .05; ***p* < .01; ****p* < .001

| | Model 1 | | Model 2 | | Model 3 | | Model 4 | |
|---|---|---|---|---|---|---|---|---|
| Predictor | $\beta$ | *p* | $\beta$ | *p* | $\beta$ | *p* | $\beta$ | *p* |
| IL-6 | -0.162 | 0.001** | -0.143 | 0.008** | -0.106 | 0.03* | -0.120 | 0.03* |
| BMI | | | -0.001 | 0.716 | -0.002 | 0.545 | -0.002 | 0.619 |
| Number of antidepressant(s) | | | -0.039 | 0.114 | -0.031 | 0.173 | -0.031 | 0.182 |
| Number of comorbidities | | | -0.020 | 0.447 | -0.016 | 0.527 | -0.016 | 0.526 |
| Number of non-psychotropic drug(s) | | | 0.020 | 0.275 | 0.017 | 0.326 | 0.016 | 0.359 |
| Sex (MALE) | | | | | 0.099 | *** | 0.095 | 0.001** |
| Tobacco (CURRENT) | | | | | -0.012 | 0.738 | -0.011 | 0.764 |
| Alcohol (CURRENT) | | | | | -0.011 | 0.680 | -0.011 | 0.673 |
| Severity(HAMD-17 items) | | | | | | | 0.002 | 0.544 |

| | Model 1 | | Model 2 | | Model 3 | | Model 4 | |
|---|---|---|---|---|---|---|---|---|
| Predictor | $\beta$ | *p* | $\beta$ | *p* | $\beta$ | *p* | $\beta$ | *p* |
| NEUTROPHILS | -0.029 | 0.025* | -0.022 | 0.141 | -0.021 | 0.089 | -0.022 | 0.091 |
| BMI | | | -0.003 | 0.551 | -0.003 | 0.591 | -0.003 | 0.501 |
| Number of antidepressant(s) | | | -0.040 | 0.242 | -0.027 | 0.331 | -0.030 | 0.293 |
| Number of comorbidities | | | -0.040 | 0.388 | -0.029 | 0.426 | -0.029 | 0.433 |
| Number of non-psychotropic drug(s) | | | 0.029 | 0.315 | 0.027 | 0.233 | 0.028 | 0.226 |
| Sex (MALE) | | | | | 0.142 | *** | 0.138 | *** |
| Tobacco (CURRENT) | | | | | -0.053 | 0.457 | -0.069 | 0.385 |
| Alcohol (CURRENT) | | | | | -0.012 | 0.706 | -0.009 | 0.792 |
| Severity(HAMD-17 items) | | | | | | | 0.003 | 0.620 |

**Table S3C Results of hierarchical regression on average network connectivity (derived from HC vs hiCRP MDD NBS network) against IL-6 and Neutrophils.** [a]$\log_{10}$ pg/L; [b]x10-3 /uL; [c]body mass index, kg/m$^2$; **p* < .05; ***p* < .01; ****p* < .001

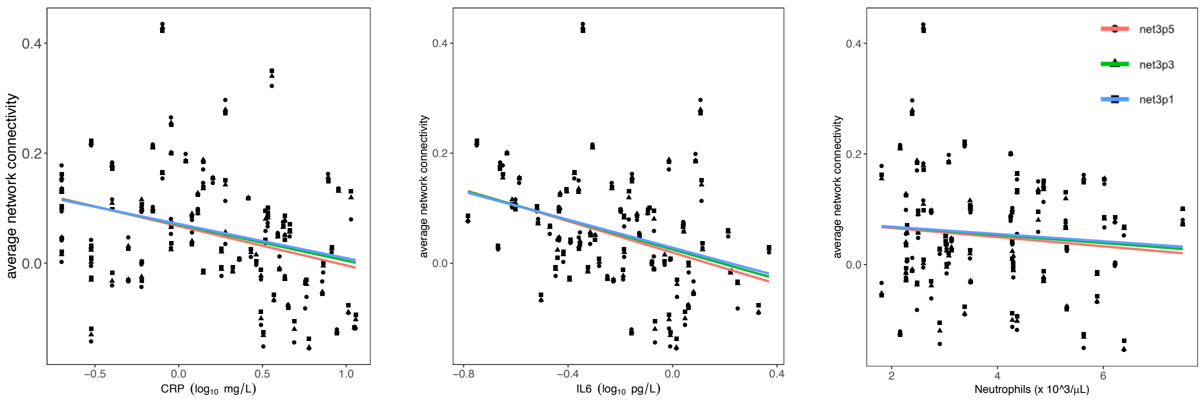


**Figure S6A Sensitivity analyses for NBS testing.** Sensitivity analyses on average network connectivity (ANC) derived from HC vs hiCRP MDD at $t_{primary} = 3.8$. Negative scaling against CRP, IL-6 and neutrophils was reproduced using average network connectivity from HC vs hiCRP MDD NBS network at 3 other $t_{primary}$ = 3.1, 3.3, 3.5. This shows that relationship observed between the two variables is not specific to threshold selected, rather an inherent feature of the yielded case-control network.

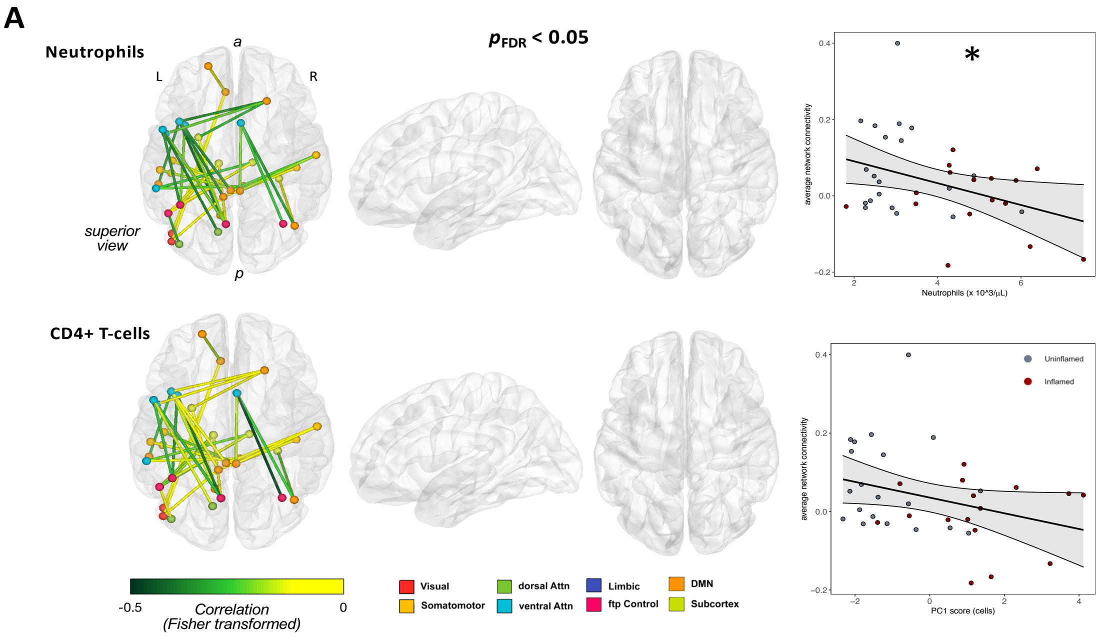


**Figure S7 Association between immune cells and FC**. **First column:** Edges implicated in CRP and IL-6 correlation with edge-wise FC were also noted here to have negative correlation with neutrophils. Weaker edge-wise correlation was observed with CD4+ T-cells. **Second column:** none survived FDR correction. **Third column:** identical association was observed between average network connectivity and neutrophils (R=-0.34, $P$=0.025, $P_{FDR}$=0.05), and CD4+ T-cells (R=-0.08, $P$=0.272) (see Table S3C).
