## Supplemental Appendix (SA) for "Dysconnectivity of a brain functional network was associated with blood inflammatory markers in depression"

### Supplemental Appendices


| | | |
|---|---|---|
| **A1** | Detailed participant inclusion and exclusion criteria | **2** |
| **A2** | Assessments performed for enrollment, stratification and for use in analyses | **3** |
| **A3** | MDD participant HAM-D score breakdown | **4** |
| **A4** | MDD antidepressant treatment resistance (ATRQ) and minor diagnostic comorbidities summary | **5** |
| **A5** | Psychological evaluation and behavioral assessment | **8** |
| **A6** | Wellcome Trust NIMA Consortium members | **10** |
| | References | **12** |

### A1 Participant inclusion and exclusion criteria

| Group | Inclusion Criteria | Exclusion Criteria |
|---|---|---|
| **All participants** | • Able and willing to provide informed consent<br>• Proficiency in English (spoken, written and comprehension)<br>• Aged 25-50 years (inclusive)<br>• Body mass index (BMI) $\leq$ 36 kg/m$^2$<br>• Able and willing to fast for 8 hours prior to blood specimen collection<br>• Willing to abstain from strenuous exercise for 72 hours prior to assessment | • Lifetime history or comorbid medical disorders that are likely to compromise immune profile (including, but not limited to, immunological disorders, cardiovascular disorders, malignancies, acute and chronic infections)<br>• Medication (in healthy controls) and concomitant therapeutics (non-antidepressant in clinical respondents) that may alter immune profile (including, but not limited to, corticosteroids)<br>• Active substance abuse or dependence within the last 6 months prior to screening<br>• Contraindications to MRI<br>• Pregnancy or breastfeeding<br>• Participation in clinical drug trial within the last 12 months prior to screening |
| **Healthy Controls** | | • Current or past history of major psychiatric disorder as defined by the DSM-V<br>• Treatment with a monoaminergic antidepressant for depressive symptoms or any other indication<br>• Baseline serum/plasma high-sensitivity CRP $\leq$ 3mg/L |
| **MDD Cases** | • Meet the DSM-V criteria for major depressive disorder (MDD) operationalized by the Structured Clinical Interview for DSM-V (SCID)<br>• Score at least 13 (from first 17 items) on the observer-rated Hamilton Rating Scale for Depression (HAM-D)<br>• Baseline serum/plasma high-sensitivity CRP $\leq$ 3mg/L for "loCRP MDD" arm<br>• Baseline serum/plasma high-sensitivity CRP $>$ 3mg/L for "hiCRP MDD" arm | • Lifetime history of bipolar disorder or other non-affective psychotic disorders |

Medications for other medical conditions (minor diagnostic comorbidities) were allowed as dictated by the patients' treating physicians, although patients were required to be medically stable as determined by medical history, physical exam and laboratory testing (see Section A4). CRP was assessed over multiple screening visits spaced 1–3 weeks apart. MDD participants with evidence of active infections (indicated by extreme CRP concentration and medical history) were excluded. Three hiCRP MDD participants with CRP > 10 mg/L (range; 10.2 – 11.4 mg/L) were included in our analyses as evidence of ongoing acute infection could not be confidently established. CRP concentration for implicated participants were stably high (within  20% or ± 2mg/L) on MRI appointment and preceding screening visit.

### A2 Assessments performed for enrollment, stratification and for use in analyses

| Evaluation | Part 1 Assessment | Part 2 Assessment |
|---|---|---|
| Visit type | First / Eligibility | Follow-up / MRI / PET |
| ***Clinical*** | | |
| Informed Consent | X | X |
| Adverse events | | X |
| Demographics / Participant details | X | X |
| Medical History | X | X |
| Family History | X | X |
| Concomitant medication | X | X |
| Antidepressant Treatment History | X | X |
| SCID (2-item and Full Section) | X | X |
| Age, Height, Weight, BMI | X | X |
| Drugs and Alcohol | X | X |
| ***Behavioral instruments*** | | |
| Hamilton depression rating scale (HAM-D) | X | X |
| Childhood Trauma Questionnaire (CTQ) | | X |
| Beck Depression Inventory II (BDI-II) | | X |
| State-Trait Anxiety Inventory (STAI) | | X |
| Chalder Fatigue Scale (CFS) | | X |
| Snaith-Hamilton Pleasure Scale (SHAPS) | | X |
| Perceived Stress Scale (PSS) | | X |
| Life Events Questionnaire (LEQ) | | X |
| ***Biological specimen sampling*** | | |
| Biomarker blood samples collection[a] | X | X |
| Blood samples for clinical laboratory[b] | X | X |
| Blood sample for genetics | | X |
| Instruction Cortisol Collection | | X |
| CSF sample collection (optional) | | X |
| ***Neuroimaging*** | | |
| MRI (sMRI, fMRI) | | X |
| $^{11}$C PK 11195 PET-MR (optional) | | X |

[a] up to 90ml of blood was collected for biomarker assessment. Initial hs-CRP assay (Part 1 assessment) was performed at on-site laboratories across the 5 recruitment centers. Part 2 hs-CRP assay was performed at a central laboratory (see S3. Biomarker Assessment), [b]15ml of blood was collected for routine blood test.

### A3 MDD participant HAM-D score breakdown

| Group / Eligibility criterion | loCRP MDD (N = 50) | | | | hiCRP MDD (N = 33) | | | |
|---|---|---|---|---|---|---|---|---|
| **SCID 2-item Screening** | **Current depression + current anhedonia** | **Only current depression** | **Only current anhedonia** | **Neither / Incomplete** | **Current depression + current anhedonia** | **Only current depression** | **Only current anhedonia** | **Neither / Incomplete** |
| N | 40 | 8 | 0 | 1 / 1 | 27 | 6 | 0 | 0 |
| **Severity Index (N)** | | | | | | | | |
| HAM-D > 17 | 19 | 4 | 0 | 0 | 15 | 3 | 0 | 0 |
| HAM-D 14-17 | 21 | 4 | 0 | 1 / 1 | 12 | 3 | 0 | 0 |
| **SCID Full Mood Disorder Assessment** | **Current episode depressed** | **Current episode HCI depressed** | **Not currently depressed** | **Diagnosis not available** | **Current episode depressed** | **Current episode HCI depressed** | **Not currently depressed** | **Diagnosis not available** |
| N | 33 | 7 | 2 | 8 | 21 | 3 | 1 | 8 |
| **Severity Index (N)** | | | | | | | | |
| HAM-D > 17 | 18 | 3 | 1 | 1 | 14 | 0 | 0 | 5 |
| HAM-D 14-17 | 15 | 4 | 1 | 7 | 7 | 3 | 1 | 3 |

Final MDD cohort for analyses (N=83) i.e. after imaging QC. Structured Clinical Interview for DSM-V (SCID-V) (1) 2-item screening was performed first to assess core symptoms of depression (low mood and anhedonia), before further assessment to establish current episode MDD diagnoses operationalised by SCID for DSM-V. HAM-D cutoff in our study global score HAM-D >13, correspond to “moderate depression” (HAM-D 14-17) and “severe depression” (HAM-D >17).

### A4 Participant medical history summary

### Antidepressant Treatment Resistance Questionnaire (ATRQ) summary

| | loCRP MDD (N=50) | | hiCRP MDD (N=33) | | |
|---|---|---|---|---|---|
| | Mean | SD | Mean | SD | p-value |
| Total number of antidepressants taken | 2.59 | 1.51 | 2.93 | 1.99 | 0.43 |
| Number of antidepressants taken with < 50% improvement response | 1.64 | 1.61 | 1.80 | 1.52 | 0.67 |
| Rate of treatment failure (%) | 60.61 | 40.92 | 61.29 | 40.25 | 0.94 |
| **Treatment-Resistance Categorisation** | | | | | |
| No (HAMD>13 - ≤17, currently not on antidepressant) (N) | 13 | | 4 | | |
| Yes (HAMD>13, on antidepressant) (N) | 28 | | 22 | | |
| Untreated (HAMD>17, currently not on antidepressant)(N) | 9 | | 7 | | |

Within final MDD cohort for analyses (N=83), no significant difference in treatment resistance was observed between loCRP MDD (N=50) and hiCRP MDD (N=33), although hiCRP MDD subgroup on average recorded lower treatment response for antidepressants i.e. greater number of antidepressants taken with < 50% improvement in symptoms.

### Psychotropics summary

| | hiCRP MDD (N=33) | loCRP MDD (N=50) |
|---|---|---|
| | *N* | *N* |
| Untreated | 11 | 22 |
| Currently not on antidepressant | 5 | 19 |
| Treatment-naive | 6 | 3 |
| **Selective Serotonin Reuptake Inhibitor (SSRI)** | | |
| Citalopram | 6 | 6 |
| Escitalopram | 0 | 3 |
| Sertraline | 6 | 7 |
| Fluoxetine | 3 | 1 |
| **Tricylic** | | |
| Nortriptyline | 1 | 0 |
| **Selective Norepinephrine Reuptake Inhibitor (SNRI)** | | |
| Duloxetine | 0 | 3 |
| Venlafaxine | 3 | 3 |
| **Norepinephrine & Specific Serotonergic Antidepressant (NaSSA)** | | |
| Mirtazapine | 2 | 4 |
| **Norepinephrine & Dopamine Reuptake Inhibitor (NDRI)** | | |
| Buproprion | 1 | 1 |

### Current and recent minor diagnostic comorbidities summary

| | hiCRP MDD (N=33) | loCRP MDD (N=50) | Healthy Control (N=46) |
|---|---|---|---|
| | *N* | *N* | *N* |
| None | 15 | 22 | 41 |
| **Disease / Disorder class(es)** | | | |
| Attention Deficit Hyperactivity Disorder | 1 | 0 | 0 |
| Adrenal | | | |
| Addison's disease | 0 | 1 | 0 |
| Anxiety | 4 | 2 | |
| Cardiovascular | | | |
| High blood pressure | 5 | 7 | 0 |
| Dermatological | | | |
| Eczema | 2 | 2 | 0 |
| Gastrointestinal | | | |
| Celiac disease | 0 | 1 | 0 |
| Parasite infection (giardia) | 0 | 1 | 0 |
| Irritable bowel syndrome | 2 | 4 | 0 |
| Gynecological | | | |
| Bacterial vaginosis (recurring) | 0 | 1 | 0 |
| Hematological | | | |
| Anemia | 1 | 0 | |
| Factor V Leiden deficiency | 0 | 1 | 0 |
| Hepatological | | | |
| Gilbert's syndrome | 0 | 0 | 1 |
| Insomnia | 3 | 4 | 2 |
| Musculoskeletal | | | |
| Sciatica_back pain | 3 | 5 | 0 |
| Neurological | | | |
| Migraine | 2 | 1 | 0 |
| Urogenital | | | |
| Painful bladder syndrome | 0 | 1 | 0 |
| Respiratory | | | |
| Asthma | 2 | 7 | 1 |
| Chest infection | 0 | 0 | 1 |
| Endocrine | | | |
| Hyperthyroidism | 0 | 1 | 0 |
| Hypothyroidism | 4 | 3 | 0 |

Chest infection episode in healthy control (N=1) was experienced 3 weeks prior to MRI appointment. Participant was prescribed one-week dose of antibiotics (indicated in non-psychotropics summary below) for week prior to eligibility assessment (one week before MRI appointment). ***recent** *medical event was defined as medical issues experienced 6-8 weeks prior to MRI appointment.*

### Non-psychotropics summary

| | hiCRP MDD (N=33) | loCRP MDD (N=50) | Healthy Control (N=46) |
|---|---|---|---|
| | *N* | *N* | *N* |
| None | 18 | 27 | 43 |
| **Drug class(es)** | | | |
| Adrenergic bronchodialator | 0 | 4 | 0 |
| Analgesic | 0 | 1 | 0 |
| Angiotensin receptor blocker | 1 | 0 | 0 |
| Antibiotic | 0 | 0 | 1 |
| Anticonvulsant | 4 | 1 | 0 |
| Antiemetic | 0 | 1 | 0 |
| Antihistamine | 0 | 2 | 1 |
| Antimicrobial | 0 | 1 | 0 |
| Antiparasitic | 0 | 1 | 0 |
| Antispasmodic | 1 | 0 | 0 |
| Antithyroid | 0 | 1 | 0 |
| Anxiolytic | 3 | 3 | 0 |
| Beta-blocker | 1 | 0 | 0 |
| Benzodiazepine | 1 | 0 | 0 |
| Calcium-channel blocker | 0 | 3 | 0 |
| Corticosteroid | 1 | 1 | 0 |
| Diuretic | 0 | 1 | 0 |
| Hormone (thyroxine) | 5 | 3 | 0 |
| Iron | 0 | 1 | 0 |
| Nonbenzodiazepine hypnotic | 0 | 0 | 1 |
| Noradrenaline reuptake inhibitor | 1 | 0 | 0 |
| NSAID | 0 | 1 | 0 |
| Opiate | 3 | 2 | 0 |
| Proton-pump inhibitor | 0 | 1 | 0 |
| Sedative-hypnotics | 0 | 1 | 0 |
| Statin | 2 | 1 | 0 |
| Triptan | 1 | 1 | 0 |

### A5 Psychological evaluation and behavioral assessment

Diagnoses of current MDD episode was operationalized by the SCID-V (1) conducted by trained non-clinician research assistants. The 2-item screening questionnaire for low mood (depression; current 2 weeks) and anhedonia (current 2 weeks) was first administered before further assessment with the full Mood Disorders SCID-I section to ascertain current MDD episode in participants. We supplemented SCID outcome with the HAM-D assessment also administered by trained non-clinician research assistants to index severity of current MDD episode in MDD cases. We used SCID and HAM-D outcomes following Part 2 assessment to ensure eligibility criteria were still met at MRI. Previously eligible MDD cases that no longer satisfied 2-item screening questionnaire and HAM-D cutoff were excluded from analyses (**see A3**).

All participants completed the following self-report standardized instruments (see footnotes Table1 and below): Beck's Depression Inventory version 2 (BDI-II), Snaith-Hamilton Pleasure Scale (SHAPS), State- Trait Anxiety Inventory (STAI), Chalder Fatigue Score (CFS), Childhood Trauma Questionnaire (CTQ), Perceived Stress Scale (PSS) and Life Events Questionnaire (LEQ).

***Clinical evaluation.*** Age, gender, medical history, and family history were documented using semi-structured clinical interview.

***Structured Clinical Interview (DSM-5 version) (1)*** Axis I Disorders (SCID-I) The 2-item screening of core depressive symptoms (low mood or depression and anhedonia) and assignment of MDD diagnosis was operationalized using SCID for Diagnostic and Statistical Manual for Mental Disorders, 5th. edition (DSM-V) Axis I Mood Disorders. The instrument was administered by trained non-clinician research assistants over two occasions i.e. during initial visit as a constituent of the Part 1 Assessment for allocation into study arm, and again during the second MRI visit as a constituent of the Part 2 Assessment, for follow-up or current evaluation (**Figure S1**). SCID is widely considered as the diagnostic gold standard for psychiatric evaluation.

***Hamilton Rating Scale for Depression (HAM-D)*** (2, 3). The HAM-D is a 21-item observer-rated instrument designed to measure severity of depression. Only the first 17 items are weighted towards severity measurement. The additional 4 items - assessing for diurnality in symptom presentation, paranoia and obsessive-compulsive behaviors - reflect the type of depression and presentation of rarely occurring symptoms, as opposed to measuring intensity of depression. Thus, the final 4 variables are excluded from the rating scale. Each item is assessed against a 5-point (0-4; 8 items) or 3-point (0-2; 9 items) scale, with reverse-scoring for item-17(Insight). Total scores range from 0-52, with greater scores indicating more severe depression. The questionnaire is extensively used and has high reliability and validity (4–6). In our study, the HAM-D was used to complement formal diagnoses of MDD (alongside SCID for DSM-V) as part of the eligibility criteria (see above) and measure severity of current depressive state in participants. HAM-D cutoff (baseline score > 13) corresponding to moderate depression and beyond was implemented based on previous literature.

***Beck Depression Inventory version 2 (BDI-II)*** ((7, 8). The 21-item self-reported questionnaire BDI-II was used to measure severity of disease. Akin to the HAM-D, the instrument indicates severity of depression through general assessment of somatic, affective and cognitive symptoms. Each item is scored against a 4-point scale (0-3), with greater total scores (ranging from 0 to 63) indicating greater severity.

***Snaith-Hamilton Pleasure Scale (SHAPS)*** (9). This 14-item self-reported instrument was used to measure hedonic tone or more strictly, anhedonia – one of the core symptoms of depression. Each item has a 4-level Likert scale i.e. “Definitely agree”, “Agree”, “Disagree” and “Strongly disagree”. The former two “Agree” responses receive a score of 1, whilst the latter two “Disagree” responses receive 0. Thus, total scores range from 0 to 14, with higher scores indicating higher levels of current anhedonia or reduced hedonic experience.

***State-Trait Anxiety Inventory (STAI)*** (10). The 40-item STAI was used to measure State-anxiety (STAI-S) and Trait-anxiety (STAI-T) with 20 items loading onto each factor (or subset) i.e. first 20 items for STAI-S and, subsequent 20 items for STAI-T. State-anxiety is a measure of the current state of anxiety i.e. severity of anxious symptom experienced by the subject. These are transient feelings. Trait-anxiety in contrast, is a persistent or consistent experience of anxiety, may be conceptualized as a personality trait and thus a risk factor for depression. Similar to the SHAPS, each item within each subset have 4-category response. Scoring is reversed for the anxiety absent variables (19 out of 40 items) which broadly assess states of confidence, calmness and security (11). Total scores range between 20 and 80 for each factor, with higher scores denoting greater anxiety.

***Chalder Fatigue Scale (CFS)*** (12). Severity of fatigue was assessed using the self-reported 11-item CFS. Both physical symptoms (items 1-7; e.g. “Do you have less strength in your muscles?”) and mental symptoms (items 8-11; e.g. “Do you have problems concentrating?”) were evaluated in measuring the severity of overall fatigue. Each item is scored on a 4-level Likert scale i.e. “Less than usual”, “No more than usual”, “More than usual”, “Much more than usual”, with increasing score for each response (0-3). Higher global scores suggest greater fatigue experience.

***Childhood Trauma Questionnaire (CTQ)*** (13). The 28-item CTQ was used to measure experience of early life adversities across five subscales (emotional abuse, physical abuse, sexual abuse, physical neglect and emotional neglect), and an additional 3-item subscale (minimization and denial) to measure bias in response i.e. tendency for respondent to under-report maltreatment. Higher scores indicate a significant history of childhood trauma.

***Perceived Stress Scale (PSS)*** (14). Perception of global stress was measured using the 10-item PSS. Each item was scored on a 5-point scale (0-5), with scoring reversal for the four reverse-worded items (items 4,5,7,8). Higher scores (ranging from 0 to 40) are indicative of greater stress appraisal.

***Life Events Questionnaire (LEQ)*** (15, 16). Evaluation of threatening and stressful life events occurring 6 months preceding assessment) was performed through the 14-item LEQ. The questionnaire is an adapted version of the LTE-Q which appraises threatening events over 12 threatening categories. In the LEQ, a 13$^{th}$ item stressful or major event – “wife or partner gave birth to a child” and a general 14$^{th}$ item – “any other significant event” was added. The instrument comprised a binary scale (“yes” or “no”) for each item and a corresponding self-appraised rating to indicate likelihood of the recent life event still affecting the respondent. Number of events was summed to produce LEQ score (frequency of recent stressors) and corresponding rating were summed to obtain global LEQ rating (severity of recent stressors). Higher prescribed ratings denote acute impact of the occurrence on respondent.

### A6 NIMA Consortium members during the sample collection and data analysis period for the BioDep Study

<u>Brighton & Sussex University Hospitals NHS Trust</u>
Dominika Wlazly

<u>Cambridgeshire & Peterborough NHS Foundation Trust</u>
Amber Dickinson, Andy Foster, Clare Knight

<u>Cardiff University</u>
Claire Leckey, Paul Morgan, Angharad Morgan, Caroline O'Hagan, Samuel Touchard

<u>GSK</u>
Shahid Khan, Phil Murphy, Christine Parker, Jai Patel, Jill Richardson

<u>Janssen</u>
Paul Acton, Nigel Austin, Anindya Bhattacharya, Nick Carruthers, Peter de Boer, Wayne Drevets, John Isaac, Declan Jones, John Kemp, Hartmuth Kolb, Jeff Nye, Gayle Wittenberg

<u>Kings College London</u>
Gareth Barker, Anna Bogdanova, Heidi Byrom, Diana Cash, Annamaria Cattaneo, Daniela Enache, Tony Gee, Caitlin Hastings, Melisa Kose, Giulia Lombardo, Nicole Mariani, Anna McLaughlin, Valeria Mondelli, Maria Nettis, Naghmeh Nikkheslat, Carmine Pariante, Karen Randall, Julia Schubert, Luca Sforzini, Hannah Sheridan, Camilla Simmons, Nisha Singh, Federico Turkheimer, Vicky Van Loo, Mattia Veronese, Marta Vicente Rodriguez, Toby Wood, Courtney Worrell, Zuzanna Zajkowska

<u>Lundbeck</u>
Brian Campbell, Jan Egebjerg, Hans Eriksson, Francois Gastambide, Karen Husted Adams, Ross Jeggo, Thomas Moeller, Bob Nelson, Niels Plath, Christian Thomsen, Jan Torleif Pederson, Stevin Zorn

<u>NHS Greater Glasgow and Clyde</u>
Catherine Deith, Scott Farmer, John McClean, Andrew McPherson, Nagore Penandes, Paul Scouller, Murray Sutherland

<u>Oxford Health NHS Foundation Trust</u>
Mary Jane Attenburrow, Jithen Benjamin, Helen Jones, Fran Mada, Akintayo Oladejo, Katy Smith

<u>Pfizer</u>
Rita Balice-Gordon, Brendon Binneman, James Duerr, Terence Fullerton, Veeru Goli, Zoe Hughes, Justin Piro, Tarek Samad, Jonathan Sporn

<u>Sussex Partnership NHS Foundation Trust</u>
Liz Hoskins, Charmaine Kohn, Lauren Wilcock

<u>University of Cambridge</u>
Franklin Aigbirhio, Junaid Bhatti, Ed Bullmore, Sam Chamberlain, Marta Correia, Anna Crofts, Tim Fryer, Martin Graves, Alex Hatton, Manfred Kitzbichler, Mary-Ellen Lynall, Christina Maurice, Ciara O'Donnell, Linda Pointon, Peter St George Hyslop, Lorinda Turner, Petra Vertes, Barry Widmer, Guy Williams

University of Glasgow
Jonathan Cavanagh, Alison McColl, Robin Shaw

University of Groningen
Erik Boddeke

University of Oxford
Alison Baird, Stuart Clare, Phil Cowen, I-Shu (Dante) Huang, Sam Hurley, Simon Lovestone, Alejo Nevado-Holgado, Elena Ribe, Anviti Vyas, Laura Winchester

University of Southampton
Madeleine Cleal, Diego Gomez-Nicola, Renzo Mancuso, Hugh Perry

University of Sussex
Mara Cercignani, Charlotte Clarke, Alessandro Colasanti, Neil Harrison, Rosemary Murray

University of Texas
Jason O'Connor

University of Toronto
Howard Mount
